# Multi-ancestry meta-analyses of lung cancer in the Million Veteran Program reveal novel risk loci and elucidate smoking-independent genetic risk

**DOI:** 10.1101/2024.04.25.24306313

**Authors:** Bryan R. Gorman, Sun-Gou Ji, Michael Francis, Anoop K. Sendamarai, Yunling Shi, Poornima Devineni, Uma Saxena, Elizabeth Partan, Andrea K. DeVito, Jinyoung Byun, Younghun Han, Xiangjun Xiao, Don D. Sin, Wim Timens, Jennifer Moser, Sumitra Muralidhar, Rachel Ramoni, Rayjean J. Hung, James D. McKay, Yohan Bossé, Ryan Sun, Christopher I. Amos, VA Million Veteran Program, Saiju Pyarajan

**Affiliations:** Center for Data and Computational Sciences (C-DACS), VA Boston Healthcare System, Boston, MA, USA; Booz Allen Hamilton, McLean, VA, USA; The University of British Columbia Centre for Heart Lung Innovation, St Paul’s Hospital, Vancouver, British Columbia, Canada; University Medical Centre Groningen, GRIAC (Groningen Research Institute for Asthma and COPD), University of Groningen, Groningen, Netherlands; Department of Pathology & Medical Biology, University Medical Centre Groningen, University of Groningen, Groningen, Netherlands; Office of Research and Development, Department of Veterans Affairs, Washington, DC, USA; Lunenfeld-Tanenbaum Research Institute, Sinai Health System, University of Toronto, Toronto, Ontario, Canada; Section of Genetics, International Agency for Research on Cancer, World Health Organization, Lyon, France; Institut universitaire de cardiologie et de pneumologie de Québec, Department of Molecular Medicine, Laval University, Quebec City, Quebec, Canada; Department of Biostatistics, University of Texas MD Anderson Cancer Center, Houston, TX, USA; Institute for Clinical and Translational Research, Baylor College of Medicine, Houston, TX, USA; Department of Medicine, Section of Epidemiology and Population Sciences, Baylor College of Medicine, Houston, TX, USA; Dan L Duncan Comprehensive Cancer Center, Baylor College of Medicine, Houston, TX, USA; Department of Medicine, Brigham and Women’s Hospital, Harvard Medical School, Boston, MA, USA

## Abstract

Lung cancer remains the leading cause of cancer mortality, despite declines in smoking rates. Previous lung cancer genome-wide association studies (GWAS) have identified numerous loci, but separating the genetic risks of lung cancer and smoking behavioral susceptibility remains challenging. We performed multi-ancestry GWAS meta-analyses of lung cancer using the Million Veteran Program (MVP) cohort and a previous study of European-ancestry individuals, comprising 42,102 cases and 181,270 controls, followed by replication in an independent cohort of 19,404 cases and 17,378 controls. We further performed conditional meta-analyses on cigarettes per day and identified two novel, replicated loci, including the 19p13.11 pleiotropic cancer locus in LUSC. Overall, we report twelve novel risk loci for overall lung cancer, lung adenocarcinoma (LUAD), and squamous cell lung carcinoma (LUSC), nine of which were externally replicated. Finally, we performed phenome-wide association studies (PheWAS) on polygenic risk scores (PRS) for lung cancer, with and without conditioning on smoking. The unconditioned lung cancer PRS was associated with smoking status in controls, illustrating reduced predictive utility in non-smokers. Additionally, our PRS demonstrates smoking-independent pleiotropy of lung cancer risk across neoplasms and metabolic traits.

## Introduction

Lung cancer remains the leading cause of overall cancer mortality, as the most prevalent cancer type in men, and the second highest in women after breast cancer^1–3^. Despite declines in smoking rates in the US since the 1980s^4^, tobacco use is currently implicated in upwards of 80% of lung cancer diagnoses^1^. Even in those who have never smoked, nor had meaningful exposure to environmental carcinogens^1,5^, there exists a heritable risk component of lung cancer conferred by genetic factors^6–8^. Differentiating the mutations which directly predispose an individual to lung cancer from those whose effect is mediated through environmental components remains challenging.

Genome-wide association studies (GWAS) have identified lung cancer risk variants associated with oncogenic processes such as immune response^7^, cell cycle regulation^9^, and those affecting DNA damage response and genomic stability^8^. Several lung cancer GWAS have also reported strong effects of genes such as *CHRNA* nicotine receptor genes which putatively increase the risk of lung cancer through behavioral predisposition towards smoking^5^. Characteristic molecular markers and genetic risk factors in smokers and never-smokers have been identified^10,11^, though fewer variants have been found in GWAS performed exclusively in never-smokers^12^.

Lung cancer has a heterogeneous genetic architecture across ancestral groups^13,14^. In the two most well-studied ancestries, European (EA) and East Asian (EAS), the majority of genome-wide significant loci are not shared^15,16^; this is in agreement with molecular studies showing differences in tumor characteristics between EA and EAS^17^. Smaller African ancestry (AA) cohorts have replicated known loci from EA or EAS^8,18^, though no AA-specific GWAS loci have been reported.

In this study, we examined lung cancer genetic variation in EA as well as in the largest AA cohort to-date. Our discovery analysis is performed in an older cohort of mostly male US veterans in the Department of Veterans Affairs Million Veteran Program (MVP)^19^. Lung cancer incidence is approximately twice as high in men than in women^2^, and additionally MVP contains a large number of cigarette smokers, positioning this biobank as particularly valuable for this analysis. We performed GWAS in overall cases of lung cancer as well as two non-small cell lung cancer (NSCLC) subtypes, adenocarcinoma (LUAD) and squamous cell lung carcinoma (LUSC).

## Results

### Genome-wide association studies for lung cancer

We performed a GWAS on overall lung cancer within EA participants in MVP (10,398 lung cancer cases and 62,708 controls; Supplementary Data 1), followed by a meta-analysis with the EA International Lung Cancer Consortium OncoArray study (ILCCO; McKay et al., 2017)^7^, for a total of 39,781 cases and 119,158 controls (Supplementary Fig. 1). The EA meta-analysis for overall lung cancer identified 26 conditionally independent SNPs within 17 genome-wide significant loci (*P*<5×10^−8^; Supplementary Fig. 2a; Supplementary Data 2). All 12 loci reported by ILCCO^7^ were confirmed, with consistent direction of effect on all single nucleotide polymorphisms (SNPs) with *P*<1×10^−5^, as well as high correlation of effect sizes and allele frequency (Supplementary Fig. 3). Of the 17 genome-wide significant loci for overall lung cancer, four were novel with respect to the broader literature: neuronal growth regulator *LSAMP*, WNT signaling regulator *NMUR2*, DNA damage repair protein *XCL2*, and hedgehog signaling regulator *TULP3*, (Table 1; Supplementary Fig. 4a-d).

**Table 1:** Novel genome-wide significant loci and their respective index variants associated with lung cancer risk in European-ancestry meta-analyses from MVP and ILCCO^7^ cohorts, MVP African ancestry, multi-ancestry meta-analyses, and in European-ancestry meta-analyses after conditioning on cigarettes per day. LUAD, adenocarcinoma; LUSC, squamous cell carcinoma; EA, effect allele; NEA, non-effect allele; EAF, effect allele frequency in the given population; OR (95% CI), odds ratio and 95% confidence interval.

| Lung cancer subtype | rsID | Cytoband | Position (hg19) | Candidate gene | EA | NEA | EAF | Discovery OR (95% CI) | Discovery *P* | Replication OR (95% CI) | Replication *P* | Combined meta-analysis OR (95% CI) | Combined meta-analysis *P* |
| --- | --- | --- | --- | --- | --- | --- | --- | --- | --- | --- | --- | --- | --- |
| **Novel loci from the European ancestry GWAS meta-analysis** |  |  |  |  |  |  |  |  |  |  |  |  |  |
| Overall | rs77045810 | 1q24.2 | 168,505,017 | *XCL2* | A | C | 0.89 | 1.10 (1.07, 1.13) | 1.43×10<sup>-10</sup> | 1.07 (1.02, 1.13) | 0.0057 | 1.09 (1.07, 1.12) | 3.94×10<sup>-12</sup> |
| Overall | rs144840030 | 3q13.31 | 117,147,326 | *LSAMP* | T | G | 0.01 | 1.31 (1.19, 1.44) | 1.09×10<sup>-8</sup> | 1.07 (0.88, 1.30) | 0.49 | 1.26 (1.16, 1.37) | 5.01×10<sup>-8</sup> |
| Overall | rs62400619 | 5q33.1 | 152,343,053 | *NMUR2* | T | C | 0.68 | 1.06 (1.04, 1.08) | 6.33×10<sup>-9</sup> | 1.03 (0.99, 1.06) | 0.16 | 1.05 (1.03, 1.07) | 1.10×10<sup>-8</sup> |
| Overall | rs9988980 | 12p13.33 | 3,038,917 | *TULP3* | T | C | 0.39 | 1.05 (1.04, 1.08) | 5.34×10<sup>-8</sup> | 1.05 (1.02, 1.09) | 0.0022 | 1.05 (1.04, 1.07) | 3.72×10<sup>-10</sup> |
| LUAD | rs67824503 | 8q24.21 | 129,535,264 | *MYC* | T | C | 0.75 | 1.10 (1.07, 1.14) | 1.81×10<sup>-8</sup> | 1.11 (1.05, 1.16) | 5.05×10<sup>-5</sup> | 1.10 (1.07, 1.14) | 4.09×10<sup>-12</sup> |
| LUAD | rs11855650 | 15q23 | 70,431,773 | *TLE3* | T | G | 0.38 | 1.09 (1.06, 1.12) | 1.12×10<sup>-8</sup> | 1.12 (1.07, 1.17) | 1.22×10<sup>-7</sup> | 1.10 (1.07, 1.13) | 1.15×10<sup>-14</sup> |
| LUSC | rs36229791 | 10q24.31 | 101,991,135 | *BLOC1S2* | A | T | 0.04 | 1.27 (1.17, 1.38) | 4.04×10<sup>-8</sup> | 1.25 (1.12, 1.41) | 1.49×10<sup>-4</sup> | 1.26 (1.18, 1.35) | 2.48×10<sup>-11</sup> |
| **Novel loci from the African ancestry GWAS** |  |  |  |  |  |  |  |  |  |  |  |  |  |
| Overall | rs78994068 | 12q24.32 | 127,225,803 | *LINC00944* | C | A | 0.01 | 2.13 (1.66, 2.72) | 1.87×10<sup>-9</sup> | 1.026 (0.681, 1.548) | 0.90 | 1.76 (1.42, 2.17) | 1.81×10<sup>-7</sup> |
| **Novel loci from the multi-ancestry meta-analysis (not genome-wide significant in the European meta-analysis)** |  |  |  |  |  |  |  |  |  |  |  |  |  |
| Overall | rs329122 | 5q31.1 | 133,864,599 | *JADE2* | A | G | 0.43 | 0.95 (0.93, 0.97) | 1.12×10<sup>-8</sup> | 0.97 (0.94, 1.00) | 0.053 | 0.96 (0.94, 0.97) | 3.69×10<sup>-9</sup> |
| Overall | rs7300571 | 12q13.11 | 47,857,826 | *RPAP3* | T | C | 0.11 | 1.08 (1.05, 1.12) | 3.47×10<sup>-8</sup> | 1.07 (1.02, 1.13) | 0.0044 | 1.08 (1.06, 1.11) | 6.48×10<sup>-10</sup> |
| **Novel loci after conditioning on cigarettes per day from the European ancestry GWAS meta-analysis** |  |  |  |  |  |  |  |  |  |  |  |  |  |
| Overall | rs1124241 | 6q16.1 | 97,722,453 | *MMS22L* | A | G | 0.22 | 1.08 (1.05, 1.11) | 1.26×10<sup>-8</sup> | 1.06 (1.02, 1.11) | 0.0062 | 1.08 (1.05, 1.10) | 3.39×10<sup>-10</sup> |
| LUSC | rs61494113 | 19p13.11 | 17,401,859 | *ABHD8* | A | G | 0.29 | 1.12 (1.07, 1.16) | 4.90×10<sup>-8</sup> | 1.10 (1.03, 1.17) | 0.0031 | 1.11 (1.08, 1.15) | 6.39×10<sup>-10</sup> |

Further association tests stratified by cancer subtypes LUAD and LUSC in MVP EA (Supplementary Fig. 2bc; Supplementary Data 3-4) replicated associations reported by ILCCO^7^ (Supplementary Fig. 3) and identified additional loci. Two novel EA meta-analysis loci were identified for LUAD, proto-oncogene *MYC* and Wnt signaling inhibitor *TLE3* (Table 1; Supplementary Fig. 4e-h). For LUSC, we identified one novel locus at 10q24.31 near NFκB inhibitor *CHUK* and *BLOC1S2*. Across all subtypes for EA meta-analysis index variants, the MVP cohort had associations with *P*<0.05 in all but one in overall lung cancer, five in LUAD, including approximately nominal significance at rs67824503 (*MYC; P*=0.057), and one in LUSC (Supplementary Data 2-4).

We investigated expression quantitative trait loci (eQTL) relationships between top SNPs from the EA meta-analysis across all lung cancer GWAS in GTEx v8 Lung^20^ and the Lung eQTL Consortium^21^ (Supplementary Data 2-4). This analysis showed that the LUSC index SNP rs36229791 on 10q24.31 was associated with the mRNA expression levels of *BLOC1S2* (Fig. 1a-d), consistent with previous TWAS^22^. *BLOC1S2* is an oncogene whose gene product is associated with centrosome function; centrosomal abnormalities have previously been observed *in vitro* in LUSC^23,24^.

**Figure 1.**
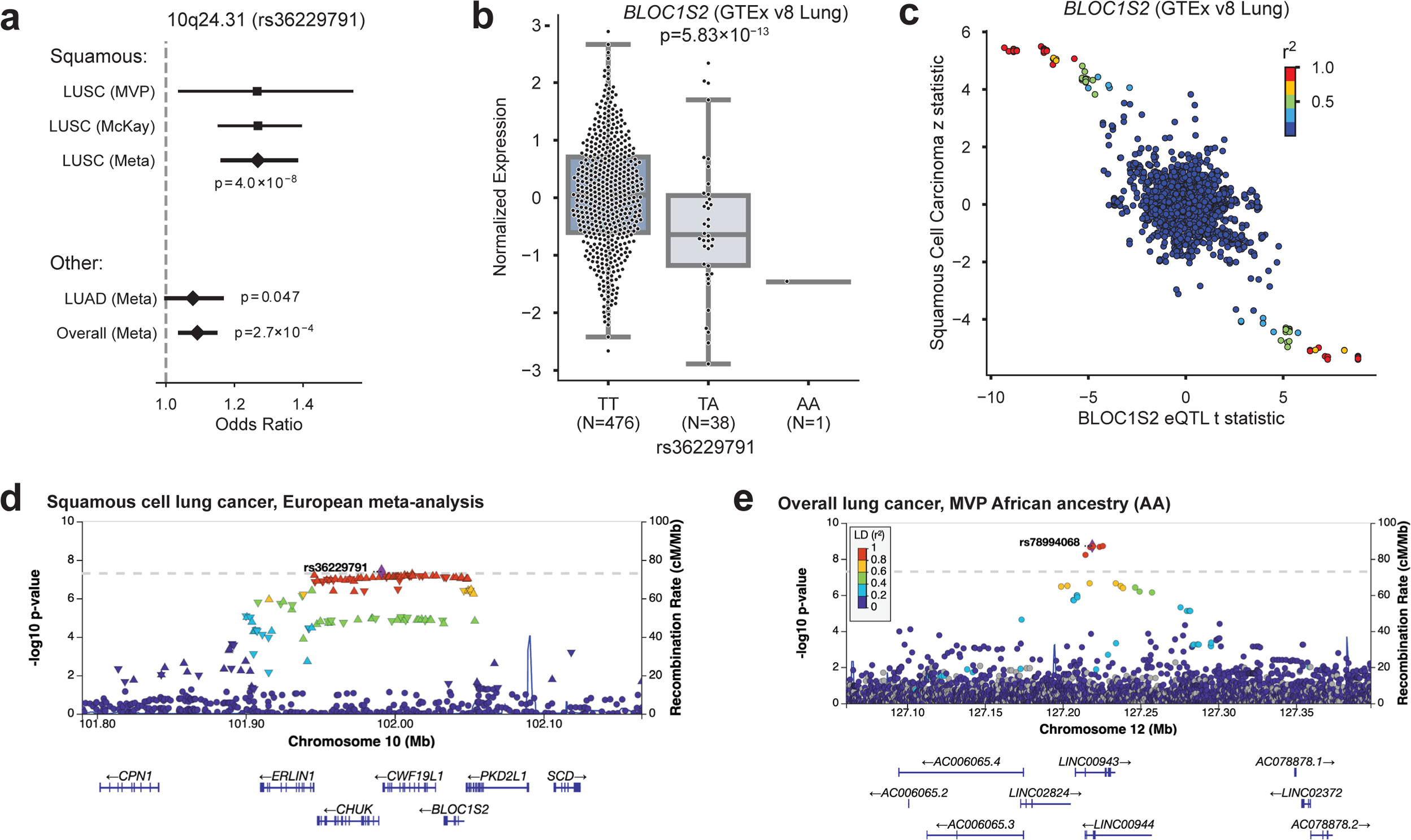
Highlighted novel GWAS loci. **a-d)** The meta-analysis of squamous cell lung carcinoma (LUSC) in European ancestry (EA) identifies a novel locus at 10q24.31. **a)** Odds ratios for rs36229791 in LUSC compared to lung adenocarcinoma (LUAD) and overall lung cancer. **b)** *BLOC1S2* expression varies by genotype at rs36229791. **c)** *BLOC1S2* eQTL t statistic vs LUSC z statistic. **d)** Regional association plot showing SNP significance and genes around lead SNP rs36229791. **e)** The African ancestry GWAS highlights a putatively novel locus on chr12 at *LINC00944*. The risk allele has effectively 0% frequency in EA.

We improved our variant selection by fine-mapping and estimating credible sets of candidate causal variants in EA meta analysis using sum of single effects (SuSiE)^25,26^ modeling. For overall lung cancer, LUAD, and LUSC, we identified 23, 23, and 9 high quality credible sets, respectively, containing 370, 246, and 192 total SNPs (Supplementary Data 5).

### GWAS in AA

We analyzed overall lung cancer risk in 2,438 cases and 62,112 controls of African ancestry (AA), the largest AA GWAS discovery cohort to date (Supplementary Fig. 5a). Two loci reached genome-wide significance in our discovery scan: 15q25, replicating the association in *CHRNA5* for AA populations reported by an earlier GWAS^18^, and a putative novel locus at 12q23 with index SNP rs78994068 (Table 1; Fig. 1e). We further performed GWAS in AA within LUAD and LUSC subtypes but found no genome-wide significant associations (Supplementary Fig. 5b-c).

The putative AA locus at 12q23 is driven by six SNPs in high linkage disequilibrium (LD; *R*^2^>0.8) found in long non-coding RNAs *LINC00943* and *LINC00944* (Fig. 1e). These imputed SNPs all had odds ratios (ORs) close to 2, with 1.3% frequency in AA and 0% in EA, consistent with gnomAD v3. *LINC00944* is highly expressed in immune cells and blood, and enriched in T cell pathways in lung tissue and cancer^27–30^. We fine-mapped this locus to define a 95% credible set (Supplementary Data 6), and annotated the functional consequence of the variants using the Variant Effect Predictor (VEP)^31^. Two variants, rs78994068 and rs115962601, were in a known enhancer regulatory region (ENSR00000974920) and thus may involve regulatory changes. However, this locus was directionally consistent but not significant in our AA replication cohort (discussed below); therefore, larger-scale AA analyses are needed to confirm this finding.

### GWAS multi-ancestry meta-analysis

We conducted a fixed-effect inverse variance-weighted multi-ancestry meta-analysis, combining the EA meta-analysis and the MVP AA GWAS for overall lung cancer, LUAD, and LUSC (Supplementary Data 7-9; Supplementary Fig. 6a-c). This analysis identified two additional novel genome-wide significant loci in overall lung cancer (Table 1; Supplementary Fig. 4i-j): ubiquitin ligase *JADE2*, previously associated with smoking initiation^32^, and RNA polymerase-associated *RPAP3*. Neither of these novel multi-ancestry meta-analysis loci were reported in a recent multi-ancestry analysis by Byun et al.^8^ that included fewer AA and more Asian ancestry samples, indicating the value our larger AA sample provided for novel discovery. All genome-wide significant EA meta-analysis associations reached genome-wide significance in the multi-ancestry meta-analysis except rs11855650 (*TLE3*) in LUAD (*P*=6.19×10^−8^). We additionally performed random effects meta-analyses using the Han-Eskin method (RE2)^33^, and observed similar *P*-values to the fixed effect meta-analysis, with all index variants *P*_RE2_<5×10^−8^ (Supplementary Data 7-9).

### Polygenic risk scoring

To gain an understanding of the penetrance and pleiotropy of lung cancer risk, we constructed PRSs based on the ILCCO summary statistics^7^ for every EA subject in MVP. As expected, the PRS was highly associated with both lung cancer risk as well as smoking behavior (Supplementary Fig. 7a-b). Even after removing individuals with any history of lung cancer risk to prevent enrichment of risk factors and comorbidities, the association with smoking behavior remained, suggesting that the PRS is partially capturing genetic smoking behavioral risk factors (Supplementary Fig. 7c). In all groups, individuals at the top decile of the PRS were at significantly higher risk of lung cancer than those in the lowest decile.

### Multi-trait conditional analysis for smoking status

Despite adjusting for smoking status, both in MVP EA and ILCCO^7^, a significant genetic correlation was observed between all subsets of lung cancer GWAS and a recently published GWAS of smoking behaviors^34^ (Fig. 2a, Supplementary Data 10). In order to remove all residual effects of smoking on lung cancer susceptibility, we conducted a multi-trait-based conditional and joint analysis (mtCOJO)^35,36^, conditioning on a GWAS for cigarettes per day^34^, which was the smoking trait most strongly correlated with overall lung cancer and subtype GWAS from the EA meta-analysis. Because lung cancer case selection also preferentially selects smokers, conventional adjustment for smoking may inadvertently cause selection bias, which functions as a collider to induce biased genetic effects^37^. mtCOJO is considered more robust to potential collider bias than conventional covariate adjustment^35,36^. The total observed-scale SNP-heritability^38^ of lung cancer risk decreased substantially after conditioning on cigarettes per day, from 5.4% to 3.1% in overall LC, from 6.7% to 5.5% in LUAD, and from 5.8% to 3.8% in LUSC (Fig. 2b; Supplementary Data 11).

**Figure 2.**
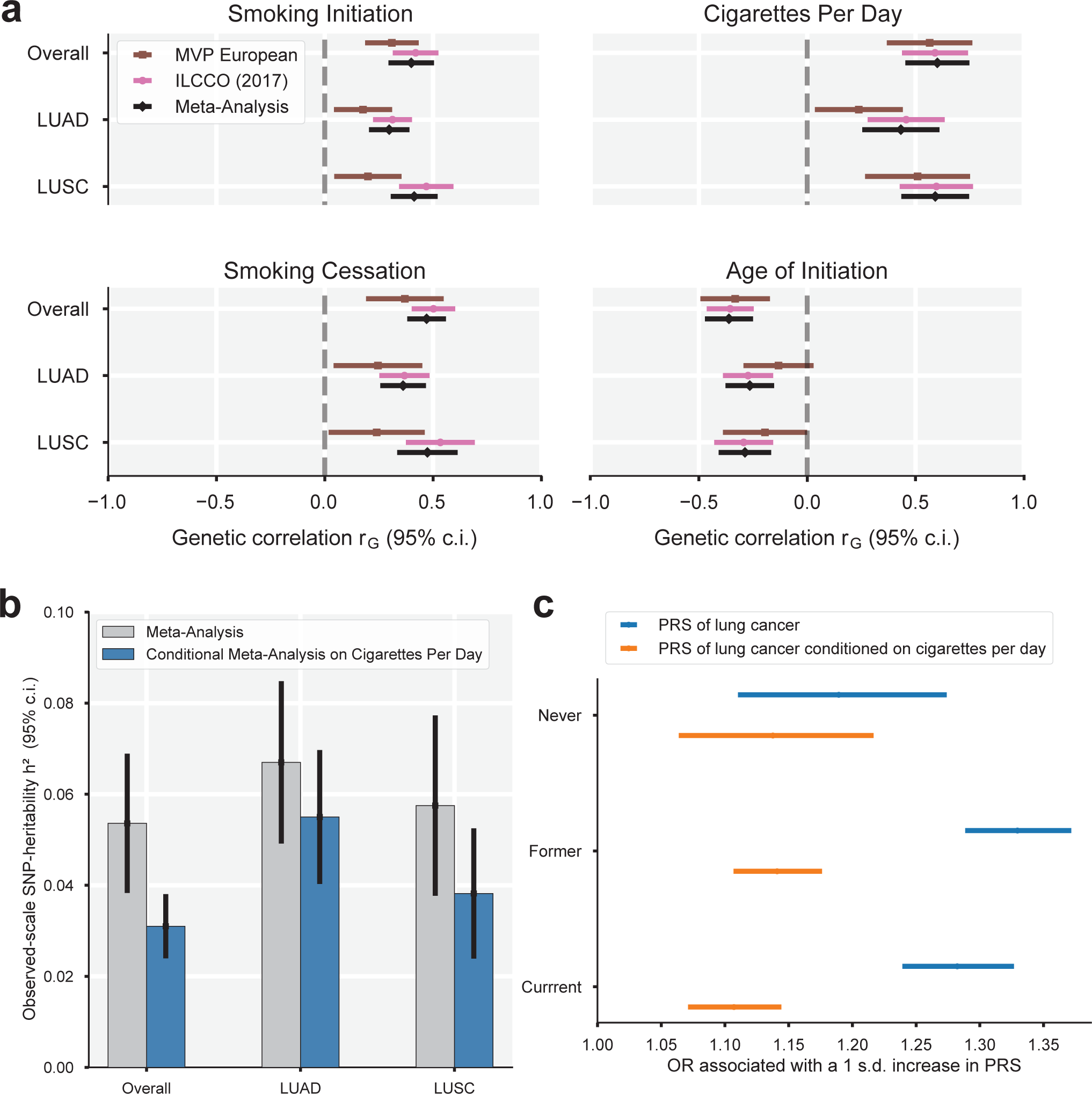
Association of lung cancer GWAS with smoking behaviors. **a)** Genetic correlations (with 95% confidence interval) between the lung cancer GWAS and smoking behaviors, including smoking initiation, cigarettes per day, smoking cessation, and age of initiation. **b)** SNP heritability for the meta-analysis and conditional meta-analysis. The heritability decreases in the conditional analysis for overall lung cancer as well as both subtypes, suggesting that some portion of the heritability of lung cancer is due to smoking behavior. **c)** Polygenic risk scores (PRS) based on standard lung cancer GWAS (blue) performs worse in never-smokers than former or current smokers, while conditioning on smoking behavior (orange) results in similar performance.

Significant loci from the conditional analyses are shown in Supplementary Fig. 8-9 and Supplementary Data 12-14. As expected, the statistical significance of loci harboring smoking-related genes (e.g., *CHRNA5*, *CYP2A6*, *CHRNA4*) dropped to below genome-wide significance after conditioning (Fig. 3). Conversely, five signals (four loci) became significant only after conditioning, including novel signals at *MMS22L* in overall lung cancer and 19p13.1 (*ABHD8*) in LUSC. *MMS22L* is a novel GWAS signal but was previously identified as overexpressed in lung cancer in genome-wide gene expression scan^39^. These may represent biological lung cancer signals partially masked by countervailing genetic effects on smoking behavior. We performed fine-mapping to identify candidate causal variants in the conditioned EA meta-analysis summary statistics, and for overall lung cancer, LUAD, and LUSC, we identified 11, 15, and 6 high quality credible sets, respectively, containing a total of 243, 277, and 78 SNPs (Supplementary Data 5).

**Figure 3.**
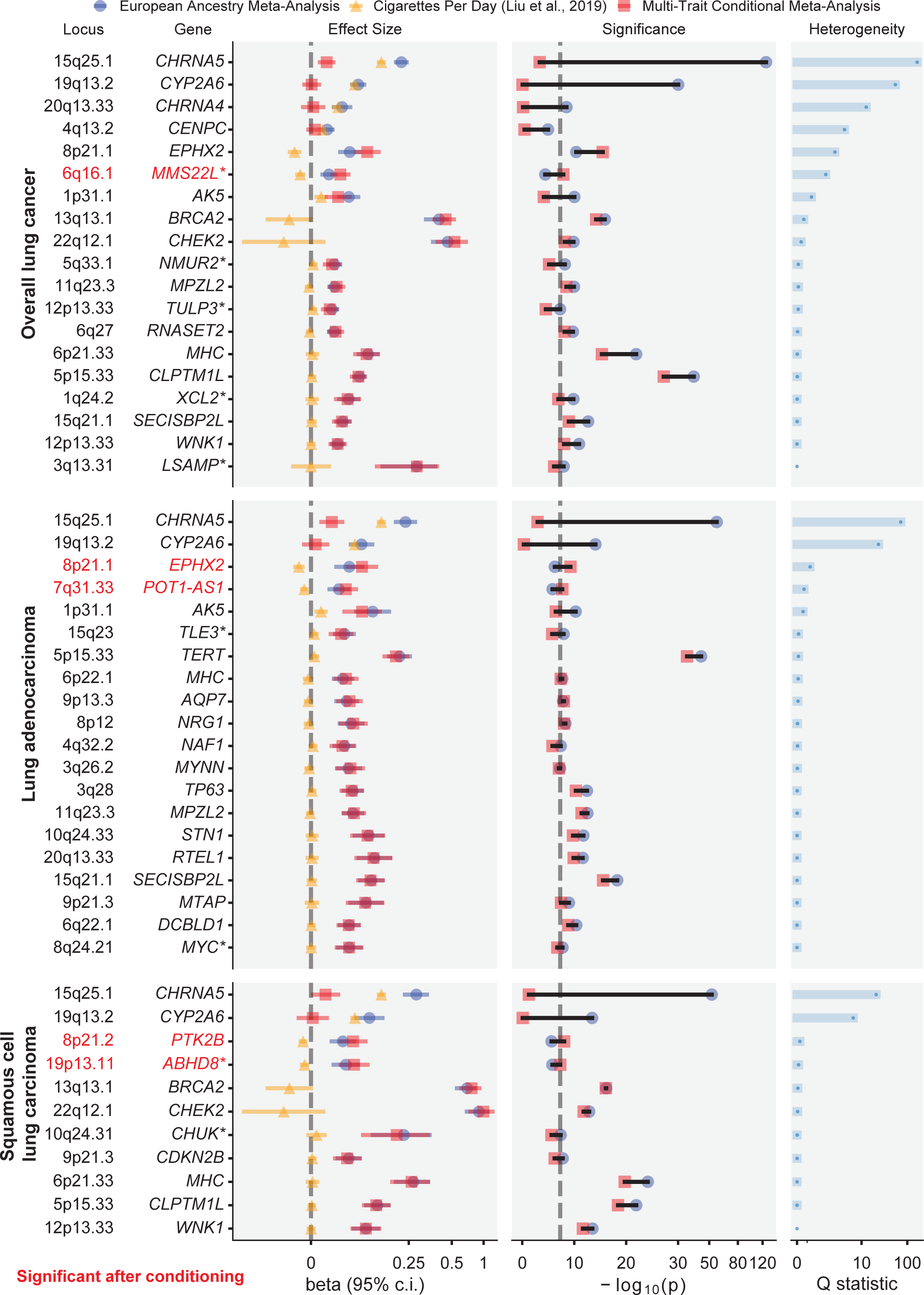
Forest plot of genome-wide significant associations. Within each cancer subtype, changes in effect size and significance are shown before and after conditioning on cigarettes per day. Novel loci are indicated by an asterisk after the gene name (*). Loci that became significant after conditioning (*P*<5×10^−8^) are in red.

We constructed PRS based on mtCOJO-conditioned ILCCO summary statistics^7^ to directly compare the predictive performance of PRS derived from the conditioned and non-conditioned GWAS in MVP EA. While the PRS based on the non-conditioned overall lung cancer GWAS exhibited reduced performance in never-smokers compared to ever-smokers, the PRS based on the conditional analysis resulted in similar performance across smoking status (Fig. 2c; Supplementary Data 15).

### Replication of novel variants in OncoArray and combined meta-analysis

We queried the OncoArray Consortium Lung Study (OncoArray) as an external non-overlapping replication dataset for our significant GWAS signals (Supplementary Data 16-17). For GWAS in EA meta-analysis for overall lung cancer, LUAD, and LUSC, we replicated five of seven novel loci (*P*<0.01) in an OncoArray European ancestry cohort: *XCL2* and *TLE3* in overall lung cancer, *MYC* and *TLE3* in LUAD, and *BLOC1S2* in LUSC. The novel African ancestry association for overall lung cancer at *LINC00944* was not replicated. We meta-analyzed OncoArray European and African ancestry participants to replicate our multi-ancestry meta-analysis signals for overall lung cancer at *RPAP3* (*P*=0.0044) and *JADE2* which bordered on nominal significance (rs329122; *P*=0.053). For the two novel loci which were identified in EA meta-analysis conditioned on cigarettes per day, we included smoking as a covariate for association analysis in the OncoArray European ancestry cohort. These association signals were replicated for overall lung cancer at *MMS22L* (*P*=0.006) and LUSC at *ABHD8* (*P*=0.003). In a variant-level replication of 137 conditionally independent discovery associations which fell within ≤1 Mb of a previously reported lung cancer GWAS signal, 134 had *P*<0.05 in OncoArray, and 42 had *P*<5×10^−8^ (Supplementary Data 18).

We then performed a combined meta-analysis of our discovery results with OncoArray replication results (Supplementary Data 18). We considered a conservative threshold of *P*=4.17×10^−9^ (*P*=5×10^−8^/12 total GWAS analyses) to be significant, which was met by 9 of the 12 loci. Because rs329122 in *JADE2* achieved the more conservative significance threshold (*P*=3.69×10^−9^), and has also been associated with smoking behavior^32^ and identified as a splicing-related variant associated with lung cancer^40^, we considered this locus to be replicated. In the combined meta-analysis we observed similar *P*-values in fixed effects and random effects (RE2) models.

Next, for all previously reported lung cancer and subtype loci in this study, we identified lung cancer associations from GWAS Catalog which fell within the same loci as our index variants (Supplementary Data 19). We confirmed two loci that previously had been reported only in a recent genome-wide association by proxy (GWAx) of lung cancer^41^: *CENPC* (rs75675343) in overall lung cancer in the EA meta-analysis (*P*=2.40×10^−8^) and the multi ancestry meta-analysis, and *TP53BP1* in overall lung cancer in the multi-ancestry meta-analysis (rs9920763; *P*=1.63×10^−8^). Our multi-ancestry meta-analysis for overall lung cancer also confirmed a recently reported locus at 4q32.2 (*NAF1*)^15^ in East Asian ancestry.

### Multi-trait analysis with breast cancer

At 19p13.1, a known pleiotropic cancer locus^42,43^, the index SNP of LUSC conditioned on smoking (rs61494113) sits in a gene-rich region where a recent fine-mapping effort of breast cancer risk loci^44^ proposed two independent associations, one affecting the regulation of *ABHD8* and *MRPL34*, and another causing a coding mutation in *ANKLE1*. Here, we used the increased power provided by a multi-trait analysis of GWAS (MTAG)^45^ of LUSC and estrogen receptor negative (ER−) breast cancer^46^ to disentangle the complex relationships between cancer risk and the genes in this locus (Fig. 4a). Overexpression of *ABHD8* has been shown to significantly reduce cell migration^42,43^. Similar odds ratios at rs61494113 were observed across LUSC and breast cancer, and MTAG enhanced the GWAS signal at this locus (Fig. 4b).

**Figure 4.**
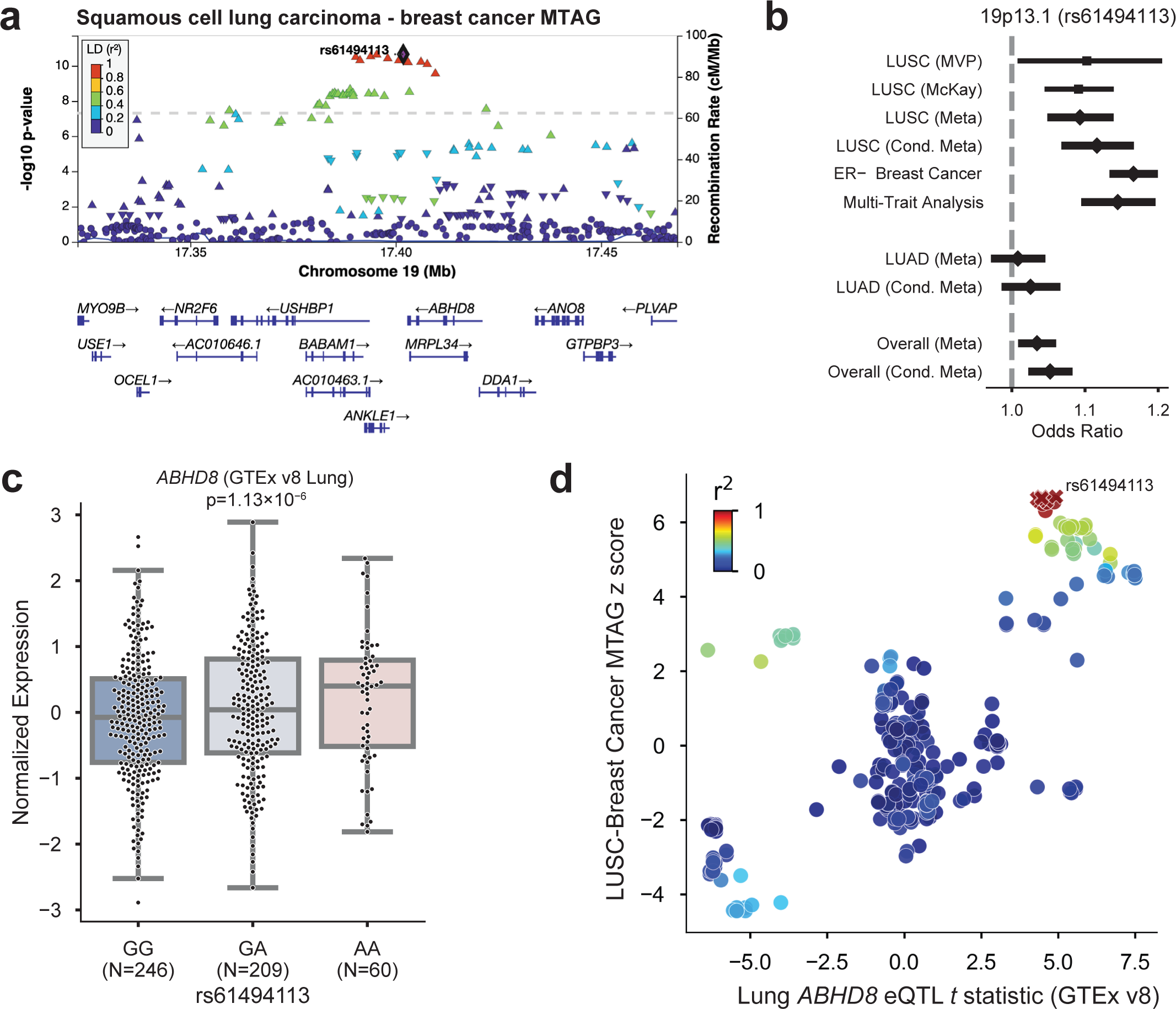
Significant locus after conditioning on smoking behavior, 19p13.11, has pleiotropic associations with ER-negative breast cancer. **a)** Regional association plot of the 19p13.11 multi-trait analysis of GWAS (MTAG) locus. **b)** Odds ratios for lead SNP rs61494113 in squamous cell lung carcinoma (LUSC), before and after conditioning, and MTAG analysis, compared to lung adenocarcinoma and overall lung cancer. c) *ABHD8* expression varies by genotype at rs61494113. d) *ABHD8* eQTL t statistic vs LUSC z statistic; red X’s indicate the 95% credible set.

We used the coloc-SuSiE method^47^ to assess colocalized associations between pairs of credible sets in this locus underlying the risk of LUSC and ER− breast cancer, allowing for multiple causal signals. We found evidence for a shared causal signal between credible sets in the LUSC conditional meta-analysis and ER− breast cancer (97.7% posterior probability; Supplementary Data 20). The index SNPs for the credible sets of LUSC conditioned on smoking and ER− breast cancer (rs61494113 and rs56069439, respectively) have *r*^2^=0.99.

The eQTL effect of *ABHD8* was replicated in multiple tissues of GTEx v8, including Lung (Fig. 4c). Interestingly, the group of SNPs in the LUSC-BC credible set did not have the most significant eQTL effect, suggesting a complex relationship between the multiple causal variants at the locus and gene expression (Fig. 4d). For instance, a recent splice variant analysis^48^ implicated splicing of *BABAM1* (a BRCA1-interacting protein) as a culprit of the associations observed in 19p13.1. Consistent with previous reports^42,43^, the cancer risk-increasing haplotype was correlated with increased expression of *ABHD8* and alternative splicing of *BABAM1*. However, there was no overlap between the 95% eQTL credible sets of *ABHD8* and *BABAM1*, and neither of the credible sets included rs61494113.

### Phenome-wide association study

Finally, to investigate the pleiotropy of lung cancer genetic risk in the absence of the overwhelming effect of smoking behavior, we performed PheWAS in MVP using the PRS scores constructed from the ILCCO summary statistics^7^ for overall lung cancer, both based on the standard GWAS (“unconditioned PRS”; Fig. 5a; Supplementary Data 21) and the GWAS conditioned on cigarettes per day using mtCOJO (“conditioned PRS”; Fig. 5b; Supplementary Data 22). Each PRS was tested for association with 1,772 phecode-based phenotypes. Overall, 240 phenotypes were associated with the unconditioned PRS and 112 were associated with the conditioned PRS at a Bonferroni-corrected significance threshold (*P*<0.05/1,772). Although lung cancer remained a top association with the conditioned PRS, the association with tobacco use disorder was greatly reduced, from an OR associated with a standard deviation increase in the PRS of 1.151 [1.142-1.160] (*P*=2.32×10^−237^) in the unconditioned PRS to OR=1.046 [1.038-1.053] (*P*=1.05×10^−32^) in the conditioned PRS. However, the effect on alcohol use disorder was only modestly attenuated between the unconditioned (OR=1.098 [1.089-1.108]; *P*=1.05×10^−87^) and conditioned LC (OR=1.078 [1.069-1.088], *P*=4.41×10^−60^) PRSs. Whether a role for alcohol in lung cancer exists independently of smoking is controversial^49,50^; this analysis suggests that may be the case. Other putatively smoking-related associations, such as chronic obstructive pulmonary disease, pneumonia, and peripheral vascular disease were greatly diminished with the conditioned PRS. Mood disorders, depression, and post-traumatic stress disorder, were also significantly associated with the unconditioned PRS but no longer significantly associated with the conditioned PRS, reflecting neuropsychiatric correlates of smoking behavior.

**Figure 5.**
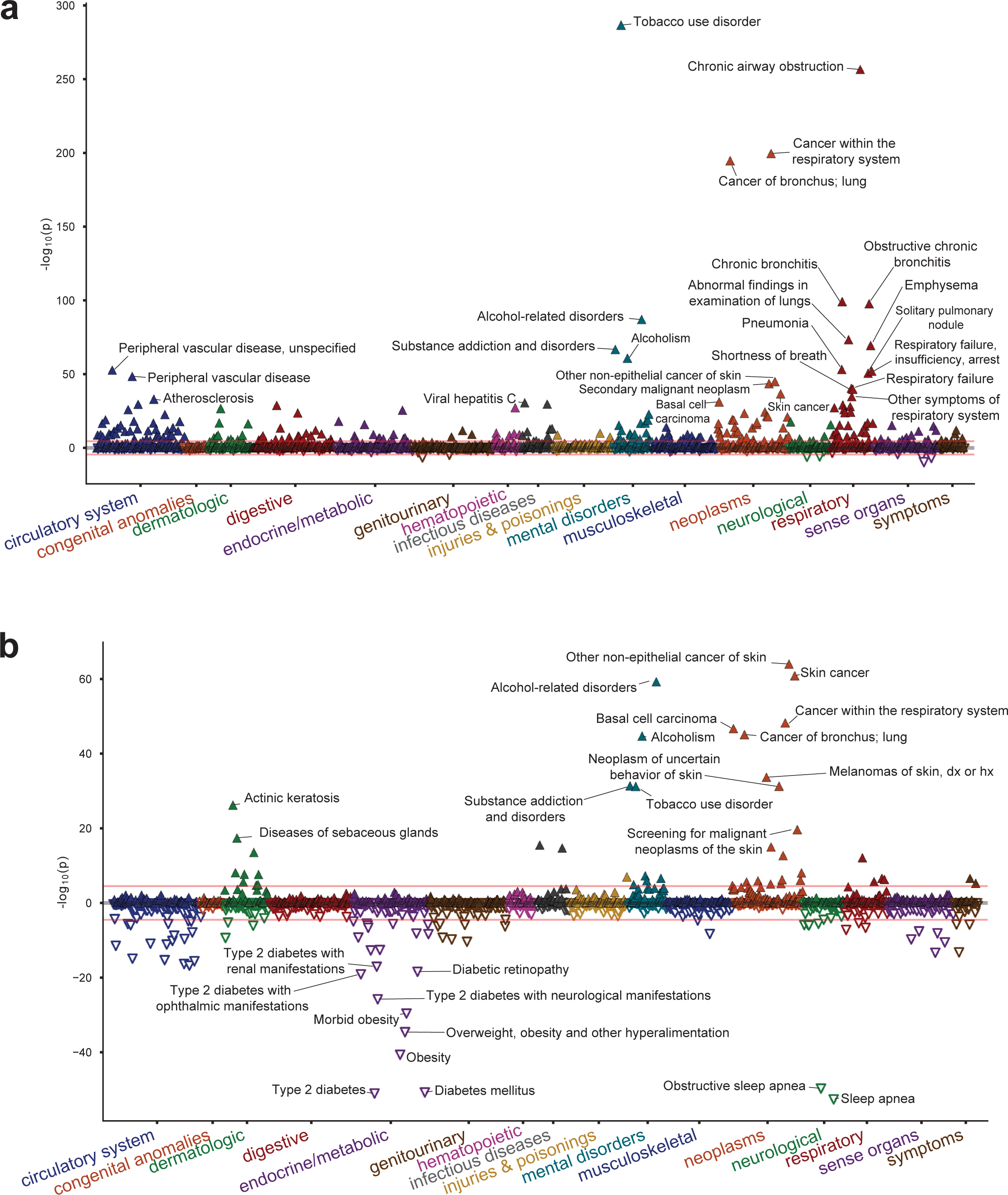
Phenome-wide association study (PheWAS) of polygenic risk scores (PRS) of lung cancer and lung cancer conditioned on cigarettes per day. **a)** PheWAS of PRS on lung cancer is mostly confounded with smoking associations. **b)** PheWAS of the conditional meta-analysis PRS shows associations with skin cancer and metabolic traits.

Intriguingly, a category of metabolic traits that were not associated with the unconditioned PRS were highly associated with the conditioned PRS and in a negative effect direction. We observed protective associations of the conditioned PRS with metabolic traits such as type 2 diabetes (OR=0.945 [0.938-0.952], *P*=9.46×10^−52^) and obesity (OR=0.952 [0.945-0.959], *P*=2.48×10^−41^). Neither were associated with the unconditioned PRS (OR=1.006 [0.999-1.014]; *P*=0.092, and OR=1.005 [0.998-1.012]; *P=*0.183, respectively). Other traits in this category included sleep apnea and hyperlipidemia. These findings are consistent with prior observational findings of an inverse relationship between BMI and lung cancer^51^ and illustrate the extent to which smoking may be a major confounder of this relationship.

Finally, we observed strong associations of the lung cancer PRS with skin cancer and related traits, such as actinic keratitis. In basal cell carcinoma, the OR increased from 1.087 [1.072-1.102] (*P*=6.06×10^−32^) with the unconditioned PRS to 1.105 [1.090-1.120] (*P*=1.82×10^−47^) with the conditioned PRS. As a sensitivity analysis, we tested the strength of this association after removing the *TERT* locus, which is prominently associated with both traits. Doing so only modestly reduced the effect of the conditioned PRS to OR=1.092 [1.077-1.107] (*P*=4.08×10^−36^). Thus, our results are consistent with a genome-wide genetic correlation between lung cancer and basal cell carcinoma that is strengthened when the effect of smoking is removed. Overall, our results suggest that the biology underlying lung cancer risk may be partially masked by the residual genetic load of smoking.

## Discussion

We identified novel lung cancer-associated loci in a new cohort of EA and AA participants, including the largest AA cohort analyzed to-date. We also show that, despite studies on the genetic basis of lung cancer risk taking smoking status into account, the effects of smoking continue to obfuscate our understanding of lung cancer genetics. In particular, we report two novel loci, at *MMS22L* (overall) and *ABHD8* (LUSC), which may be partially masked by countervailing genetic effects on smoking. Our replication analysis which adjusted for smoking pack-years confirmed these loci. Additionally, our analyses demonstrated that PRSs for lung cancer contain large uncorrected genetic loading for smoking behavioral factors. Our results indicate that controlling for these factors can improve risk assessment models, potentially improving lung cancer screening even for non-smokers. Finally, our phenomic scans comparing PRSs derived from GWAS with and without genomic conditioning on smoking showed divergent associations across numerous traits, especially metabolic phenotypes.

The increased sample size in this study enabled the interpretation of multiple causal variants underlying the gene-rich *ADHL8*-*BABAM1* region, synthesizing prior observations into a clearer understanding of this locus. Our other novel loci strengthen established lung cancer mechanisms. We identify for the first time a susceptibility locus at *MYC*, a well-known oncogene and master immune regulator. *XCL2* is involved in cellular response to inflammatory cytokines^52^. *LSAMP* is a tumor suppressor gene in osteosarcoma^53^, and 3q13.31 homozygous deletions have been implicated in tumorigenesis^54^. *TLE3* is a transcriptional corepressor involved in tumorigenesis and immune function^55^. The transcription factor *TULP3* has been implicated in pancreatic ductal adenocarcinoma and colorectal cancer^56^. *XCL2*, *NMUR2,* and *TULP3* may also be related to cancer progression via G-protein-coupled receptor (GPCR) signaling pathways^57^. *JADE2* expression has been experimentally linked to NSCLC^58^, and has been identified in GWAS of smoking behavior^34^. Finally, DNA damage repair mechanisms emerge, including *RPAP3*, an RNA polymerase that may be involved in DNA damage repair regulation^59^, and *MMS22L* which repairs double strand breaks^60^.

Although smoking is the major risk factor for lung cancer, it is important to clearly disentangle the effect of smoking to fully understand the complex genetic and environmental causes of lung cancer. Our approach enables the development of new polygenic scores, which can improve precision medicine applications for lung cancer in both smokers and nonsmokers.

## Author contributions statement

Drafted the manuscript: B.R.G., M.F., S.-G. J., A.K.S., E.P., A.K.D., S.P.

Acquired the data: B.R.G., S.-G. J., A.K.S., Y.S., P.D., U.S., D.D.S., W.T., J.M., S.M., R.R., R.J.H., J.D.M., Y.B., C.I.A., S.P.

Analyzed the data: B.R.G., S.-G. J., M.F., A.K.S., Y.S., P.D., U.S., Y.B., R.S.

Critically revised the manuscript for important intellectual content: all authors.

## Acknowledgements

This work was supported by award #MVP000 from the United States Department of Veterans Affairs (VA) Million Veteran Program. The contents of this publication are the sole responsibility of the authors and do not necessarily represent the views of VA or the United States Government. Where authors are identified as personnel of the International Agency for Research on Cancer/World Health Organization, the authors alone are responsible for the views expressed in this article, and they do not necessarily represent the decisions, policy, or views of the International Agency for Research on Cancer/World Health Organization. Full consortium acknowledgements for MVP and the ILCCO OncoArray study^7^ are provided in Supplementary Information.

## Subject terms and techniques

Biological sciences > Cancer > Lung cancer

Biological sciences > Genetics > Genetic association study > Genome-wide association studies

## Data Availability

The full summary level association data from the individual population analyses in MVP will be available upon publication via the dbGaP study accession number phs001672.

## Competing interests

S.-G.J. is an employee and shareholder of BridgeBio Pharma. The other authors declare no competing interests.

## Methods

### Cohort definition

Patients were identified from MVP participants^19^ utilizing clinical information available through the United States Department of Veterans Affairs (VA) Corporate Data Warehouse (CDW) with ICD codes for primary lung cancer. Occurrences of the ICD-9 codes 162.3, 162.4, 162.5, 162.8, and 162.9 or the ICD-10 codes C34.10, C34.11, C34.12, C34.2, C34.30, C34.31, C34.32, C34.80, C34.81, C34.82, C34.90, C34.91, and C34.92 were used in case identification. Patients with secondary lung cancer were excluded from the cohort using ICD-9/10 codes 197.x, C78.00, C78.01, and C78.02. Additional patients were identified in the VA Cancer Registry using ICD-O site, including lung/bronchus, other respiratory system or intrathoracic organs, or trachea. The Cancer Registry was also used to determine the lung cancer subtypes LUAD and LUSC among cases.

Preliminary totals of 18,633 and 10,845 patients with MVP participation were identified from the VA CDW and Cancer Registry, respectively. A combined cohort of 20,631 unique patients was generated for further analysis. The cohort was predominantly male (∼95%) with a median age of 64–68 for sub-cohorts, depending on ancestry assignments and cancer subtypes. The cohort was curated further to remove any participant with missing data. The final cohorts are described in Supplementary Data 1.

Once patients were identified from VA’s CDW and Cancer Registry, cases were used to gather records related to age, sex, smoking status, and ancestry. Smoking status included former, current, and never, based on the MVP survey at the time of enrollment and on electronic medical records. Ancestry was defined using a machine learning algorithm that harmonizes self-reported ethnicity and genetic ancestry (HARE)^61^. All analyses described here were performed on patients of EA or AA ancestry in ancestry-stratified cohorts. Additionally, the cohorts were further stratified by lung cancer subtypes for analysis. Matched controls were selected based on age, gender, smoking status, and HARE assignments. Age was binned into 5-year intervals for this purpose.

### Array genotyping, genotype quality control, and principal component analysis

Genotyping and quality control were conducted as described previously^62^. Briefly, we removed all samples with excess heterozygosity (F statistic<-0.1), excess relatedness (kinship coefficient≥0.1 with 7 or more MVP samples), and samples with call rates <98.5%. Additional samples with a mismatch between self-reported sex and genetic sex were removed.

Principal component (PC) analysis was conducted using PLINK 2.0^63^ (v2.00a3LM), on a pruned set of SNPs (window size 1Mb, step size 80, *r*^2^<0.1, minor allele frequency (MAF)<0.01, Hardy-Weinberg equilibrium *P*<1×10^−10^, missingness rate<10%) within European ancestry (EA) and African ancestry (AA) on unrelated individuals, where unrelated individuals were defined as greater than third-degree relatives as previously described^62^. PCs were then projected onto related individuals in EA.

### Imputation

Prior to imputation, a within-cohort pre-phasing procedure was applied across the whole cohort by chromosome using Eagle2^64^. Imputation was then conducted on pre-phased genotypes using Minimac4^65^ and the 1000 Genomes Phase 3 (v5) reference panel^66^ in 20Mb chunks and 3Mb flanking regions. Quality of imputation (Minimac Rsq or INFO) was then re-computed in EA and AA separately to be used as filters for respective GWAS. Imputed loci reaching genome-wide significance were tested for deviation from Hardy-Weinberg equilibrium (HWE) in 61,538 EA controls (Supplementary Data 23). Of the 93 conditionally independent SNPs across the GWAS analyses, 6 SNPs had a significant (*P <* 1×10^−6^) HWE signal; unsurprisingly, the strongest HWE signal was from SNPs in the Major Histocompatibility Complex region. However, none of the 12 novel loci reported in Table 1 significantly deviated from HWE.

### Association analyses

For the EA lung cancer overall and subtype GWAS, we performed standard logistic regression using PLINK 2.0 (v2.00a2LM)^63^ with a matched control design. EA GWAS was performed in unrelated individuals, defined as greater than third-degree relatives. For the AA lung cancer overall and subtype analyses, because the case numbers were smaller, we performed a mixed-model logistic regression using REGENIE (v1.0.6.7)^67^; REGENIE applies a whole genome regression model to control for relatedness and population structure, and includes a Firth correction to control for bias in rare SNPs as well as case-control imbalance. GWAS covariates for each ancestry included age, age-squared, sex, and smoking status as a categorical variable (current, former, never), and the first ten principal components. Participants with missing smoking status (n=786) were removed.

### EA meta-analysis

We performed inverse-variance weighted meta-analyses of MVP-EA summary statistics and summary statistics previously reported by ILCCO^7^ using METAL (v20100505)^68^ with scheme STDERR. Significant inflation across GWAS and meta-analyses was not observed (all genomic control values (λ) for GWAS in this study ≤1.15). Only variants present in both studies were meta-analyzed. We further performed a sensitivity analysis using the Han-Eskin random effects model (RE2) in METASOFT v2.0.1 ^33^.

### Lung eQTL consortium

The lung tissues used for eQTL analyses were from human subjects who underwent lung surgery at three academic sites: Laval University, University of British Columbia (UBC), and University of Groningen. Genotyping was carried out using the Illumina Human1M-Duo BeadChip. Expression profiling was performed using an Affymetrix custom array (see GEO platform GPL10379). Only samples that passed genotyping and gene expression quality controls were considered for eQTL analysis, leaving sample sizes of 409 for Laval, 287 for UBC, and 342 for Groningen. Within each set, genotypes were imputed in each cohort with the Michigan Imputation Server^65^ using the Haplotype Reference Consortium^69^ version 1 (HRC.r1-1) data as a reference set, and gene expression values were adjusted for age, sex, and smoking status. Normalized gene expression values from each set were then combined with ComBat^70^. eQTLs were calculated using a linear regression model and additive genotype effects as implemented in the Matrix eQTL package in R^71^. Cis-eQTLs were defined by a 2 Mb window, i.e., 1 Mb distance on either side of lung cancer-associated SNPs. Pre-computed lung eQTLs were also obtained from the Genotype-Tissue Expression (GTEx) Portal^20^. Lung eQTLs in GTEx (version 8) are based on 515 individuals and calculated using FastQTL^72^.

### Fine-mapping

We performed Bayesian fine-mapping the genome-wide significant loci from EA meta-analysis and AA using the FinnGen fine-mapping pipeline^73^ (https://github.com/FINNGEN/finemapping-pipeline) and SuSiE^25,26^. Pairwise SNP correlations were calculated directly from imputed dosages on European-ancestry MVP samples from this analysis using LDSTORE 2.0^73^. The maximum number of allowed causal SNPs at each locus was set to 10. Fine-mapping regions which overlapped the major histocompatibility complex (MHC; chr6:25,000,000-34,000,000) were excluded. High quality credible sets were defined as those with minimum *r*^2^<0.5 between variants. The functional consequences of the AA credible set variants were annotated using the Variant Effect Predictor (VEP)^31^.

### Replication analysis

External replication was performed for all genome-wide significant associations in overall lung cancer, LUAD, and LUSC in OncoArray Consortium Lung Study (OncoArray)^8,74^. Replication for genome-wide significant multi-ancestry associations was performed in a fixed effects meta-analysis of OncoArray CEU Europeans for significant EA meta-analysis associations, and in an YRI AA meta-analysis composed of 5 studies^8^ for significant MVP AA associations. Meta-analysis associations from this study were replicated against a meta-analysis of these OncoArray groups. To replicate significant variants from EA analysis conditioned on smoking, pack-years was additionally included as a covariate in replication cohorts. There was no participant overlap between the replication cohorts and the ILCCO study^7^ used in the discovery scan. Covariates included the first five genetic principal components and participant study sites. Proxy SNPs were used to replicate known associations at rs75675343 (rs2318539/4:67831628:C:A; R^2^ =1) and rs4586884 (rs4435699/4:164019500:C:G; R^2^ =0.999).

### Multi-ancestry meta-analysis

A multi-ancestry meta-analysis of MVP EA and AA cohorts with summary statistics previously reported by ILCCO^7^ was conducted in METAL^68^ using an inverse variance-weighted fixed effects scheme. Only variants present in two or more cohorts were meta-analyzed. Index variants were defined using the two-stage “clumping” procedure implemented in the Functional Mapping and Annotation (FUMA) platform^75^. In this process, genome-wide significant variants are collapsed into LD blocks (*r*^2^>0.6) and subsequently re-clumped to yield approximately independent (*r*^2^<0.1) signals; adjacent signals separated by <250kb are ligated to form independent loci. Novel variants are defined as meta-analysis index variants located >1Mb from previously reported lung cancer associations. We additionally performed a sensitivity analysis using the random effects model (RE2) in METASOFT v2.0.1^33^.

### Polygenic risk score (PRS) calculation

We used PRS-CS^76^ to generate effect size estimates under a Bayesian shrinkage framework, and then used PLINK 2.0 (v2.00a3LM)^63^ to linearly combine weights into a risk score using a global shrinkage prior of 1×10^−4^, which is recommended for less polygenic traits. Finally, scores were normalized to a mean of 0 and a standard deviation of 1.

### Multi-trait analyses

In order to remove all residual effects of smoking on lung cancer susceptibility, we conducted a multi-trait meta-analysis^35^ conditioned on cigarettes per day, which was shown to be most significantly correlated with all lung cancer GWAS^34^. The meta-analysis was performed on the EA meta-analysis summary statistics using mtCOJO, part of the GCTA software package^77^. An LD reference was constructed from 50,000 MVP EA samples.

Multi-trait analysis of GWAS (MTAG)^45^ (v0.9.0) was applied using genome-wide LUSC summary statistics after conditioning on cigarettes per day, and estrogen receptor negative (ER−) breast cancer summary statistics^46^ which were munged using LDSC (v1.01)^38^. Single causal variant colocalization between LUSC conditioned on cigarettes per day and ER-breast cancer was performed using Coloc (R; version 4)^78^ for variants at *ABHD8* (chr19: 17,350,000 to 17,475,000). A posterior probability > 0.9 for Hypothesis 4 (both traits are associated and share a single causal variant) was used as the criteria for colocalization.

### Heritability and genetic correlations

Linkage Disequilibrium score regression (LDSC) v1.0.1 was used to calculate observed-scale SNP-heritability^38^ using lung cancer and subtypes summary statistics, before and after conditioning on cigarettes per day. Pairwise genetic correlations were estimated between lung cancer and subtypes from MVP, ILCCO^7^, and EA meta-analysis, and four smoking traits (smoking initiation, cigarettes per day, smoking cessation, and age of initiation)^34^.

### Conditional and joint SNP analysis

To find independently associated genome-wide significant SNPs at each locus in a stepwise fashion, we used GCTA-COJO using the --cojo-slct option. An LD reference was constructed from 50,000 MVP EA samples. Variants with MAF<0.01 in the COJO reference panel were not included in identification of independent signals. LDTrait^79^ was queried to identify previously published significant GWAS variants within 1Mb of our index variants in all populations. Novel loci were defined as those at which the index variant was not within ±500_kb of previously reported genome-wide significant lead SNPs for lung cancer or its subtypes in any ancestry.

### Phenome-wide association study (PheWAS)

We conducted a PheWAS of electronic health record-derived phenotypes and lab results in EA subjects using either the normalized PRS as the predictor or independently associated genome-wide significant SNPs. Comparison of unconditioned PRS PheWAS and conditioned PRS PheWAS were based on ILCCO summary statistics^7^ and used MVP EA as the out-of-sample test set. Associations were tested using the R PheWAS package^80^ version 0.1 with QC procedures described previously^81^. Control and sex-based exclusion criteria were applied.

## Supplementary Figure captions

**Supplementary Fig. 1. Study overview.** Genome-wide association studies were performed in Million Veteran Program (MVP) European and African ancestry (AA) cohorts for overall lung cancer, adenocarcinoma, and squamous cell carcinoma. MVP and International Lung Cancer Consortium OncoArray (ILCCO) European cohorts were meta-analyzed, and further meta-analyzed with AA for multi-ancestry meta-analysis. Multi-trait conditional meta-analysis was performed on EA using average cigarettes per day from Liu et al. (2019). Replication and combined meta-analysis was performed using external OncoArray cohorts.

**Supplementary Fig. 2. Manhattan plots and quantile-quantile (QQ) plots for European meta-analyses.** Manhattan and QQ plots are shown for **a)** overall lung cancer; **b)** lung adenocarcinoma (LUAD); and **c)** squamous cell lung carcinoma (LUSC). Cytoband positions for significant loci are noted in each Manhattan plot; putatively novel loci identified in this study are in red; externally replicated novel loci are indicated by a box. Genomic control (λ) values, LDSC intercepts, and sample sizes are inset in QQ plots.

**Supplementary Fig. 3. Effect allele frequency concordance between International Lung Cancer Consortium OncoArray (ILCCO) and Million Veteran Program European ancestry (EA) GWAS. (a-c)** Effect allele frequency concordance for all variants tested in both studies with *P*<1×10^−5^ in ILCCO for **a)** overall lung cancer, **b)** lung adenocarcinoma, and **c)** squamous cell lung carcinoma. Points are styled based on significance level in MVP. **(d-f)** Effect size concordance for genome-wide significant variants in **d)** overall lung cancer, **e)** lung adenocarcinoma, and **f)** squamous cell lung carcinoma. One-to-one concordance is shown as a dashed line. Index variants from the EA meta-analysis between ILCCO and MVP are annotated by locus. Novel significant loci after meta-analysis are annotated in red.

**Supplementary Fig. 4. Genome-wide significant novel lung cancer loci.** Forest plots (left) and regional Manhattan plots (right) for novel loci from European meta-analysis: **a)** *XCL2*, **b)** *LSAMP*, **c)** *NMUR2*, **d)** *TUPL3*, **e)** *MYC*, **f)** *TLE3*, and **g)** *BLOC1S2*; and from multi-ancestry meta-analysis: **h)** *JADE2*; **i)** *RPAP3*.

**Supplementary Fig. 5. Manhattan plots and quantile-quantile (QQ) plots for MVP African ancestry.** Manhattan and QQ plots are shown for **a)** African ancestry overall lung cancer; **b)** lung adenocarcinoma (LUAD); and **c)** squamous cell lung carcinoma (LUSC). Cytoband positions for significant loci are noted in each Manhattan plot; putatively novel loci identified in this study are in red. Genomic control (λ) values and sample sizes are inset in QQ plots.

**Supplementary Fig. 6. Manhattan plots and quantile-quantile (QQ) plots for multi-ancestry meta-analyses.** Manhattan and QQ plots are shown for **a)** the multi-ancestry meta-analysis in overall lung cancer; **b)** lung adenocarcinoma (LUAD); and **c)** squamous cell lung carcinoma (LUSC). Cytoband positions for significant loci are noted in each Manhattan plot; novel loci not identified in the European meta-analysis are in red; externally replicated novel loci are indicated by a box. Genomic control (λ) values and sample sizes are inset in QQ plots.

**Supplementary Fig. 7. Association of the lung cancer polygenic risk score (PRS) with lung cancer by smoking status. a)** Association of the lung cancer PRS with overall lung cancer risk. The risk of lung cancer reached an odds ratio (OR) of 2.51 (95% confidence interval: 1.80, 3.51) in the top decile. **b)** Association of the lung cancer PRS with lung cancer risk in never-smokers. Among never-smokers, lung cancer risk reached an OR of 2.67 (2.40, 2.98) in the top decile. **c)** Association of the lung cancer PRS with lung cancer risk in ever-smokers with no history of lung cancer. The top PRS decile was associated with an OR of 1.25 (1.18, 1.32).

**Supplementary Fig. 8. Manhattan plots and quantile-quantile (QQ) plots for European meta-analyses conditioned on cigarettes per day.** Manhattan and QQ plots for **a)** overall lung cancer conditioned on cigarettes per day; **b)** lung adenocarcinoma (LUAD) conditioned on cigarettes per day; and **c)** squamous cell lung carcinoma (LUSC) conditioned on cigarettes per day. Cytoband positions for significant loci are noted in each Manhattan plot; novel loci not identified in the European meta-analysis are in red; externally replicated novel loci are indicated by a box. Genomic control (λ) values, LDSC intercepts, and sample sizes are inset in QQ plots.

**Supplementary Fig. 9. Novel loci for overall lung cancer and squamous cell carcinoma conditioned on smoking.** Forest plots (left) and regional Manhattan plots (right) for novel loci identified in the European meta-analysis conditioned on cigarettes per day: a) *MMS22L* in overall lung cancer and b) *ABHD8* in squamous cell lung cancer.

## Notes

### Author Declarations

Central IRB of the VA Office of Research & Development gave ethical approval for this work.

