## Supplementary Figures for "Multi-ancestry meta-analyses of lung cancer in the Million Veteran Program reveal novel risk loci and elucidate smoking-independent genetic risk"

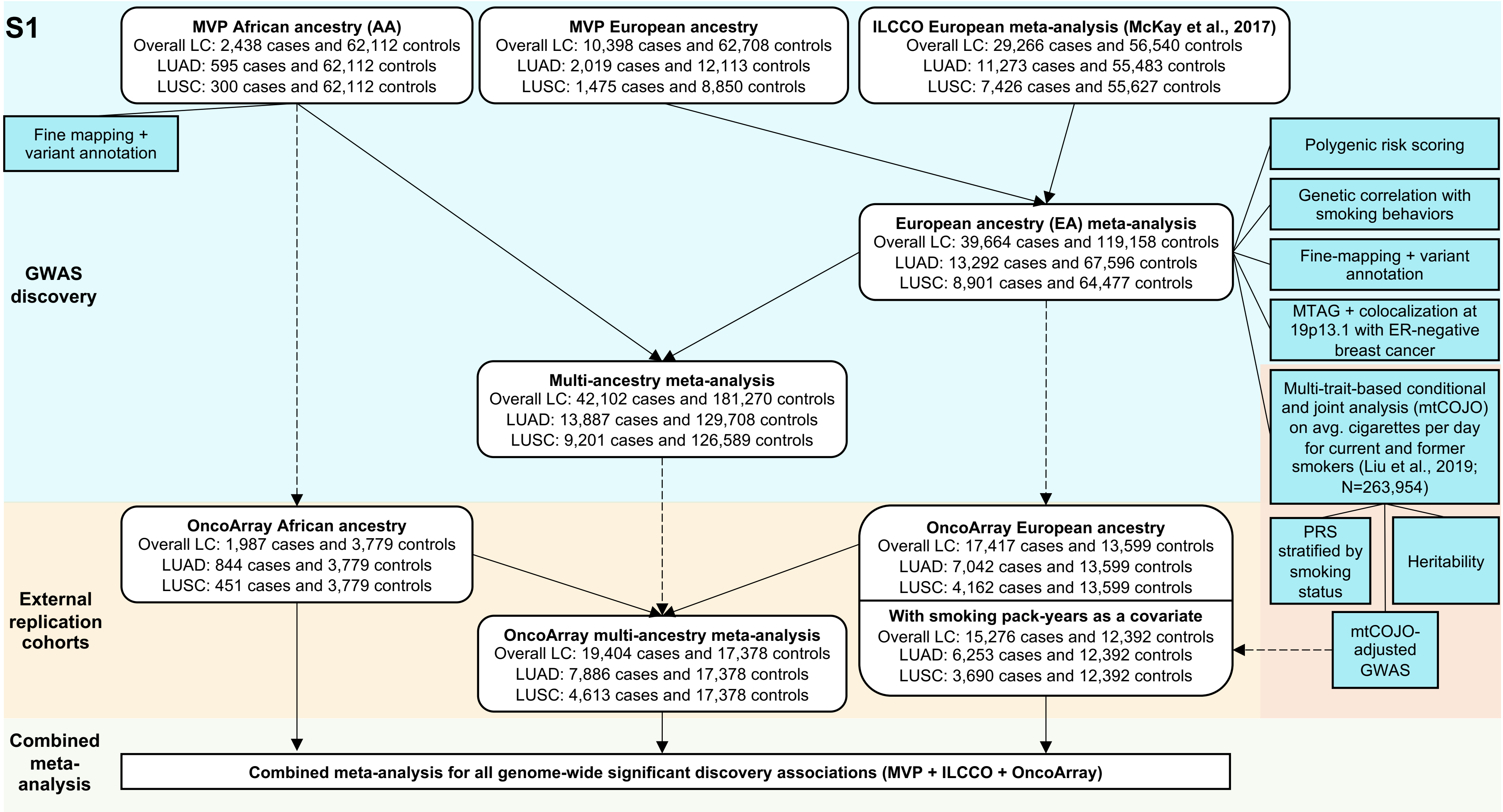


**Supplementary Fig. 1. Study overview.** Genome-wide association studies were performed in Million Veteran Program (MVP) European and African ancestry (AA) cohorts for overall lung cancer, adenocarcinoma, and squamous cell carcinoma. MVP and International Lung Cancer Consortium OncoArray (ILCCO) European cohorts were meta-analyzed, and further meta-analyzed with AA for multi-ancestry meta-analysis. Multi-trait conditional meta-analysis was performed on EA using average cigarettes per day from Liu et al. (2019). Replication and combined meta-analysis was performed using external OncoArray cohorts.

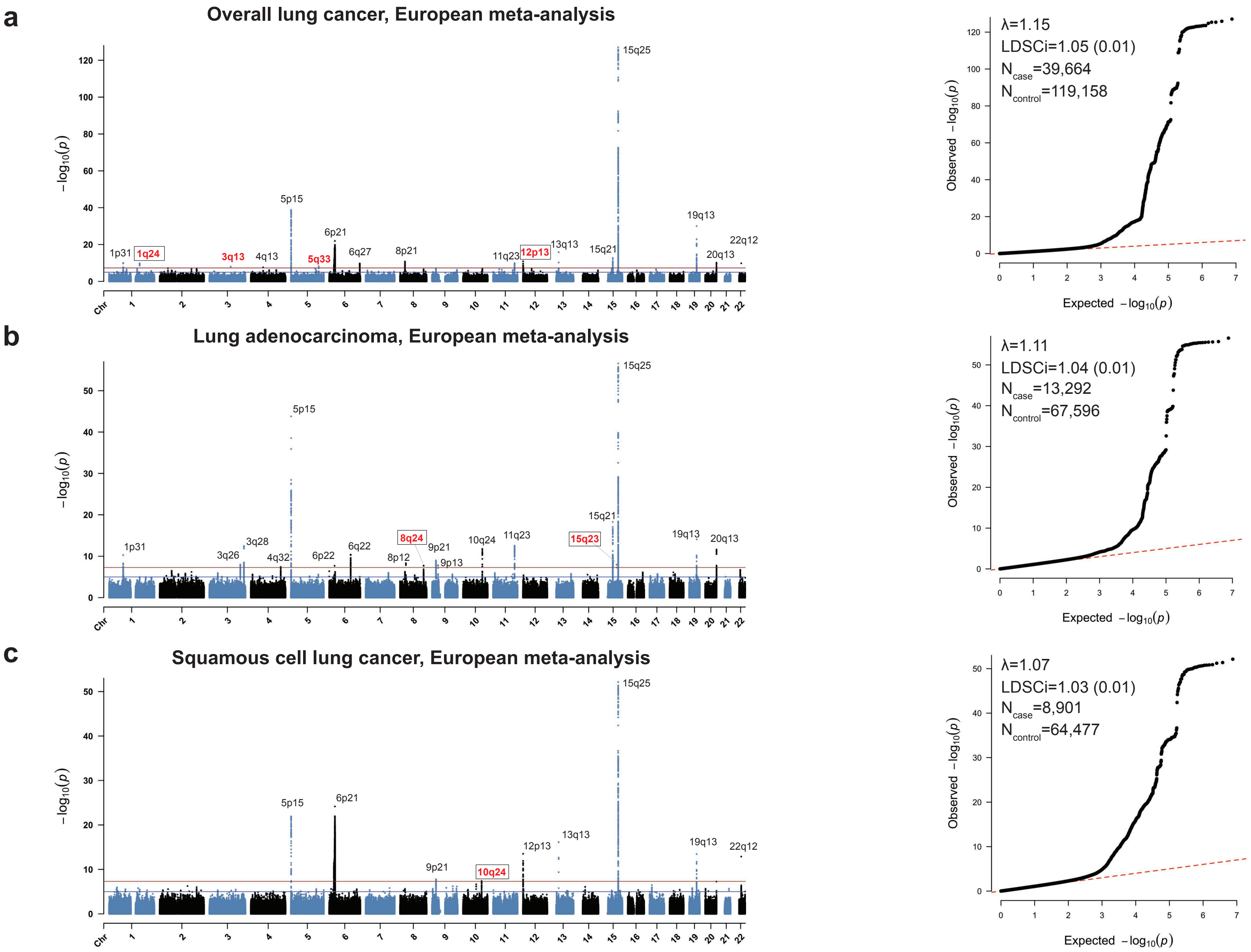


**Supplementary Fig. 2. Manhattan plots and quantile-quantile (QQ) plots for European meta-analyses.** Manhattan and QQ plots are shown for **a)** overall lung cancer; **b)** lung adenocarcinoma (LUAD); and **c)** squamous cell lung carcinoma (LUSC). Cytoband positions for significant loci are noted in each Manhattan plot; putatively novel loci identified in this study are in red; externally replicated novel loci are indicated by a box. Genomic control (λ) values, LDSC intercepts, and sample sizes are inset in QQ plots.

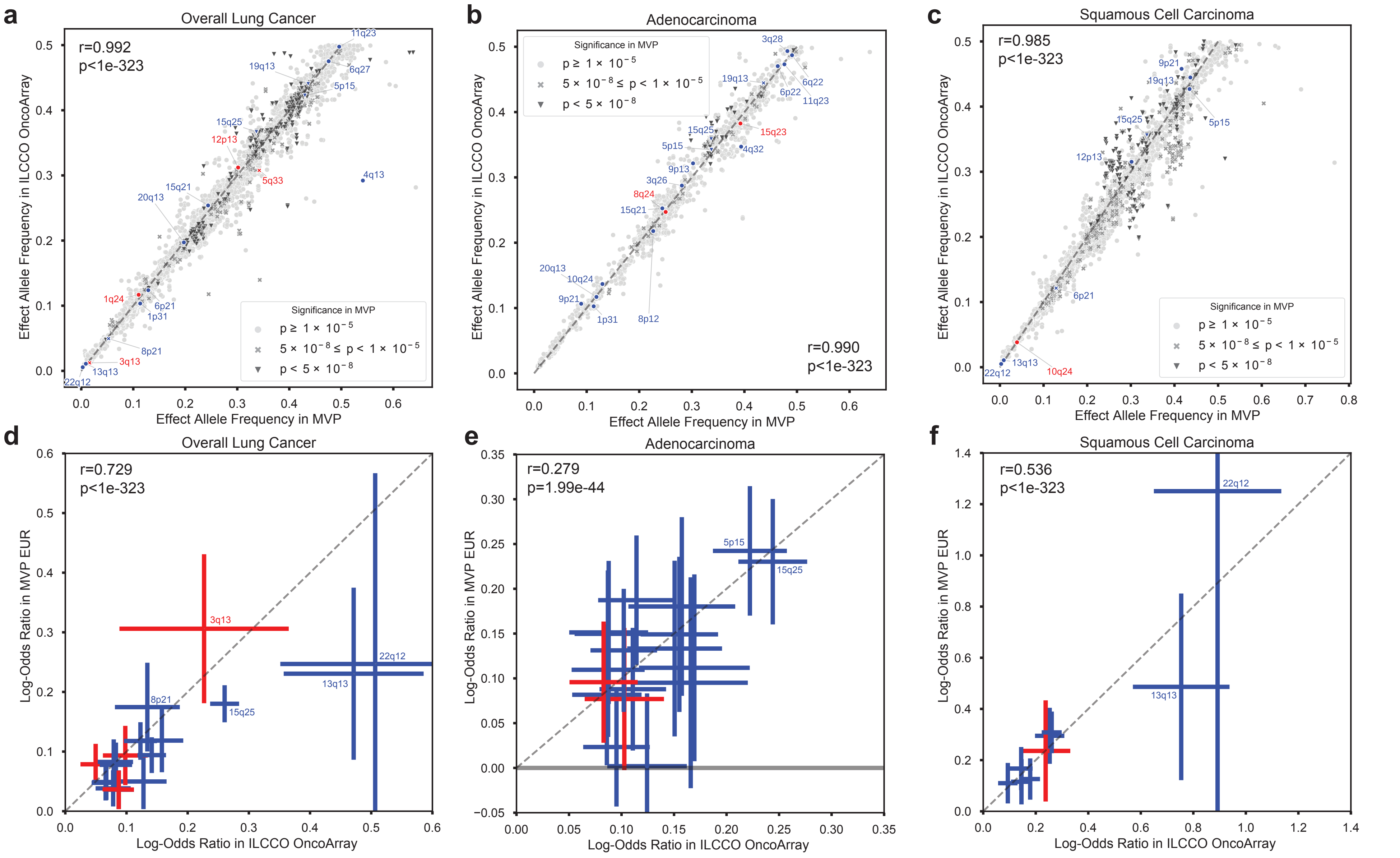


**Supplementary Fig. 3. Effect allele frequency concordance between International Lung Cancer Consortium OncoArray (ILCCO) and Million Veteran Program European ancestry (EA) GWAS. (a-c)** Effect allele frequency concordance for all variants tested in both studies with $P<1\times10^{-5}$ in ILCCO for **a)** overall lung cancer, **b)** lung adenocarcinoma, and **c)** squamous cell lung carcinoma. Points are styled based on significance level in MVP. **(d-f)** Effect size concordance for genome-wide significant variants in **d)** overall lung cancer, **e)** lung adenocarcinoma, and **f)** squamous cell lung carcinoma. One-to-one concordance is shown as a dashed line. Index variants from the EA meta-analysis between ILCCO and MVP are annotated by locus. Novel significant loci after meta-analysis are annotated in red.

**S4a**

#### Overall lung cancer, rs77045810 (XCL2; 1q24.2) : Novel and replicated

| Cohort | EA freq. | P-value | Odds ratio |
|---|---|---|---|
| MVP European | 0.89 | 2.29E-04 | 1.10 |
| ILCCO (McKay et al. 2017) | 0.88 | 1.51E-07 | 1.10 |
| European meta-analysis | 0.89 | **1.43E-10** | 1.10 |
| European meta-analysis cond. smoking | 0.89 | 1.14E-07 | 1.10 |
| OncoArray European (replication) | 0.89 | 0.006 | 1.07 |
| MVP African | 0.94 | 0.807 | 1.02 |
| OncoArray African (replication) | 0.94 | 0.512 | 0.95 |
| Multi-ancestry meta-analysis | 0.89 | 2.71E-10 | 1.10 |
| OncoArray multi-ancestry (replication) | 0.89 | 0.014 | 1.06 |
| Combined meta-analysis (European meta-analysis + OncoArray European) | 0.89 | 3.94E-12 | 1.09 |

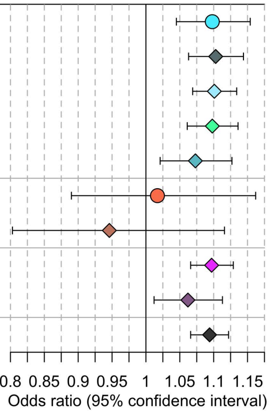


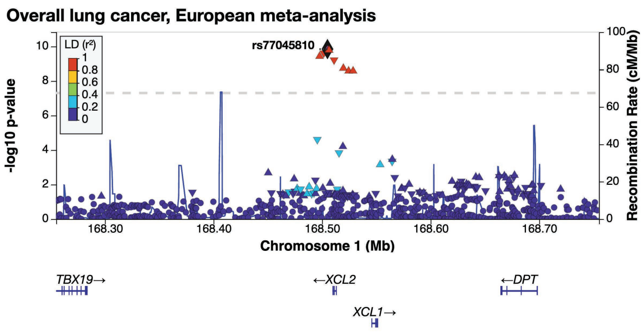


**b**

#### Overall lung cancer, rs144840030 (LSAMP; 3q13.31) : Putatively novel

| Cohort | EA freq. | P-value | Odds ratio |
|---|---|---|---|
| MVP European | 0.02 | 1.58E-06 | 1.36 |
| ILCCO (McKay et al. 2017) | 0.01 | 0.001 | 1.25 |
| European meta-analysis | 0.01 | **1.09E-08** | 1.31 |
| European meta-analysis cond. smoking | 0.01 | 7.52E-07 | 1.31 |
| OncoArray European (replication) | 0.01 | 0.487 | 1.07 |
| MVP African | 0.00 | 0.572 | 1.22 |
| OncoArray African (replication) | 0.00 | 0.760 | 1.12 |
| Multi-ancestry meta-analysis | 0.01 | 9.43E-09 | 1.31 |
| OncoArray multi-ancestry (replication) | 0.01 | 0.453 | 1.07 |
| Combined meta-analysis (European meta-analysis + OncoArray European) | 0.01 | 5.01E-08 | 1.26 |

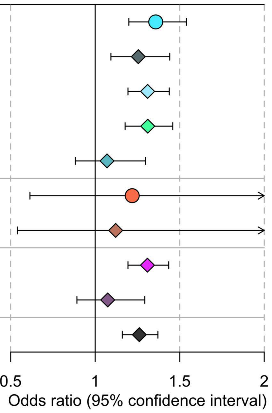


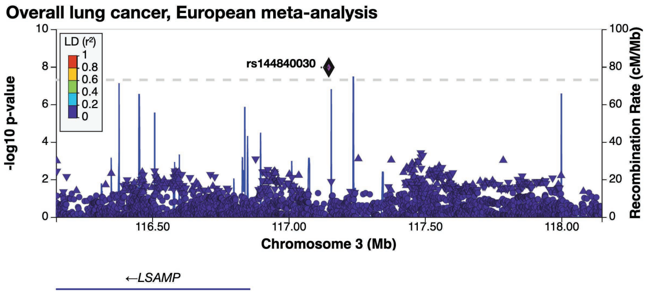


**c**

#### Overall lung cancer, rs62400619 (NMUR2; 5q33.1) : Putatively novel

| Cohort | EA freq. | P-value | Odds ratio |
|---|---|---|---|
| MVP European | 0.66 | 6.86E-06 | 1.08 |
| ILCCO (McKay et al. 2017) | 0.69 | 9.12E-05 | 1.05 |
| European meta-analysis | 0.68 | **6.32E-09** | 1.06 |
| European meta-analysis cond. smoking | 0.68 | 6.94E-06 | 1.05 |
| OncoArray European (replication) | 0.69 | 0.156 | 1.02 |
| MVP African | 0.48 | 0.974 | 1.00 |
| OncoArray African (replication) | 0.49 | 0.859 | 0.99 |
| Multi-ancestry meta-analysis | 0.66 | 2.96E-08 | 1.06 |
| OncoArray multi-ancestry (replication) | 0.66 | 0.220 | 1.02 |
| Combined meta-analysis (European meta-analysis + OncoArray European) | 0.68 | 1.10E-08 | 1.05 |

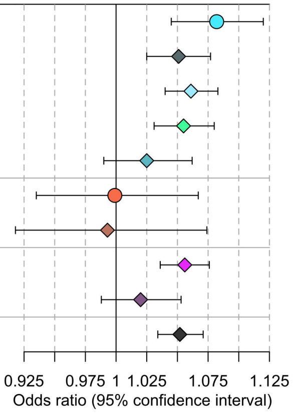


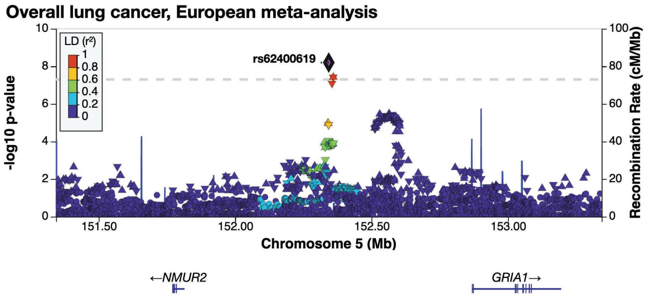

**S4d**

### Overall lung cancer, rs9988980 (TULP3; 12p13.33) : Novel and replicated

| Cohort | EA freq. | P-value | Odds ratio |
|---|---|---|---|
| MVP European | 0.38 | 5.12E-06 | 1.08 |
| ILCCO (McKay et al. 2017) | 0.40 | 6.81E-04 | 1.04 |
| European meta-analysis | 0.39 | **5.34E-08** | 1.05 |
| European meta-analysis cond. smoking | 0.39 | 2.94E-05 | 1.05 |
| OncoArray European (replication) | 0.41 | 0.002 | 1.05 |
| MVP African | 0.59 | 0.801 | 0.99 |
| Multi-ancestry meta-analysis | 0.41 | 2.77E-07 | 1.05 |
| Combined meta-analysis (European meta-analysis + OncoArray European) | 0.40 | 3.72E-10 | 1.05 |

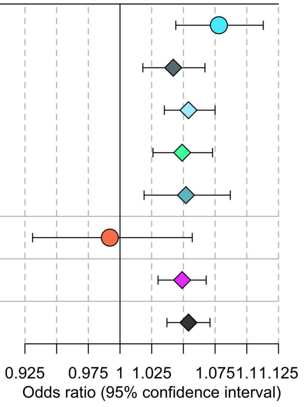


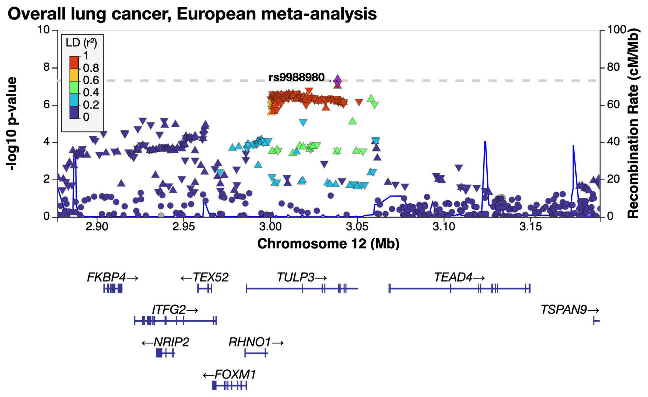


**e**

### Lung adenocarcinoma, rs67824503 (MYC; 8q24.21) : Novel and replicated

| Cohort | EA freq. | P-value | Odds ratio |
|---|---|---|---|
| MVP European | 0.75 | 0.057 | 1.08 |
| ILCCO (McKay et al. 2017) | 0.75 | 9.80E-08 | 1.11 |
| European meta-analysis | 0.75 | **1.80E-08** | 1.10 |
| European meta-analysis cond. smoking | 0.75 | 1.84E-07 | 1.10 |
| OncoArray European (replication) | 0.76 | 5.05E-05 | 1.10 |
| MVP African | 0.76 | 0.133 | 1.11 |
| OncoArray African (replication) | 0.76 | 0.028 | 1.17 |
| Multi-ancestry meta-analysis | 0.75 | 5.69E-09 | 1.10 |
| OncoArray multi-ancestry (replication) | 0.76 | 5.30E-06 | 1.11 |
| Combined meta-analysis (European meta-analysis + OncoArray European) | 0.76 | 4.09E-12 | 1.10 |

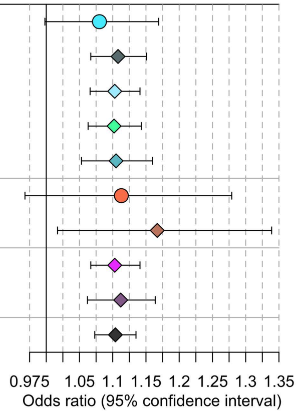


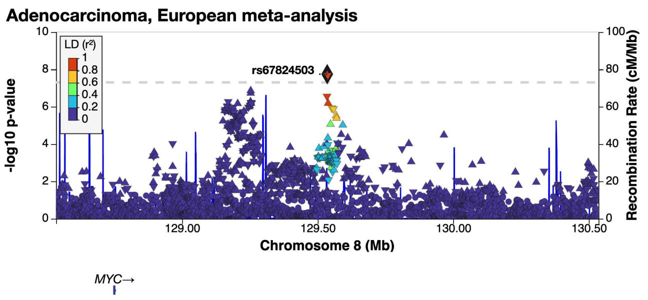


**f**

### Lung adenocarcinoma, rs11855650 (TLE3; 15q23) : Novel and replicated

| Cohort | EA freq. | P-value | Odds ratio |
|---|---|---|---|
| MVP European | 0.39 | 0.006 | 1.10 |
| ILCCO (McKay et al. 2017) | 0.38 | 5.60E-07 | 1.09 |
| European meta-analysis | 0.38 | **1.12E-08** | 1.09 |
| European meta-analysis cond. smoking | 0.38 | 1.78E-06 | 1.08 |
| OncoArray European (replication) | 0.39 | 1.22E-07 | 1.12 |
| MVP African | 0.37 | 0.544 | 0.96 |
| Multi-ancestry meta-analysis | 0.38 | 6.19E-08 | 1.08 |
| Combined meta-analysis (European meta-analysis + OncoArray European) | 0.39 | 1.15E-14 | 1.10 |

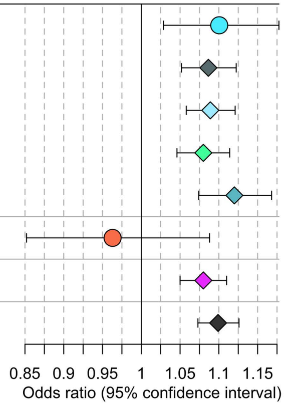


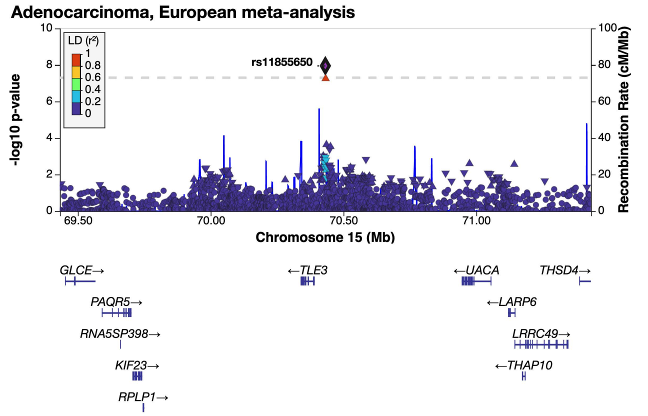

**S4g**

#### Squamous cell lung carcinoma, rs36229791 (CHUK/BLOC1S2; 10q24.31) : Novel and replicated

| Cohort | EA freq. | P-value | Odds ratio |
|---|---|---|---|
| MVP European | 0.04 | 0.019 | 1.27 |
| ILCCO (McKay et al. 2017) | 0.04 | 6.85E-07 | 1.27 |
| European meta-analysis | 0.04 | **4.04E-08** | 1.27 |
| European meta-analysis cond. smoking | 0.04 | 2.36E-06 | 1.25 |
| OncoArray European (replication) | 0.04 | 1.49E-04 | 1.25 |
| MVP African | 0.04 | 0.506 | 1.18 |
| OncoArray African (replication) | 0.04 | 0.691 | 0.92 |
| Multi-ancestry meta-analysis | 0.04 | 3.35E-08 | 1.26 |
| OncoArray multi-ancestry (replication) | 0.04 | 4.26E-04 | 1.22 |
| Combined meta-analysis (European meta-analysis + OncoArray European) | 0.04 | 2.48E-11 | 1.26 |

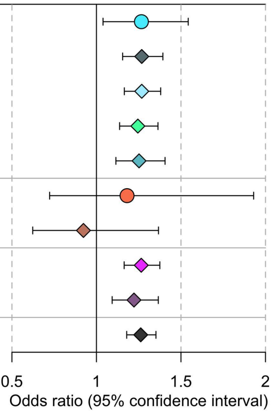


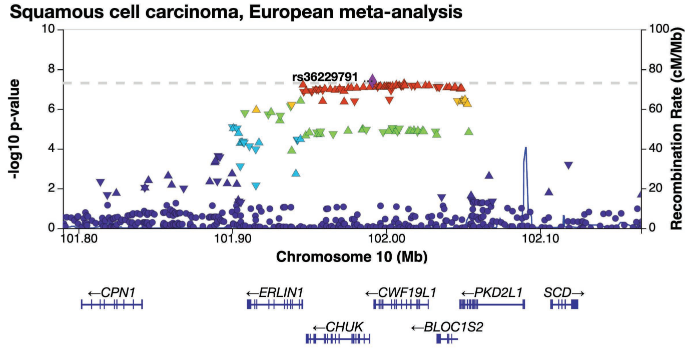


**h**

#### Overall lung cancer, rs329122 (JADE2; 5q31.1) : Novel and replicated

| Cohort | EA freq. | P-value | Odds ratio |
|---|---|---|---|
| MVP European | 0.41 | 0.002 | 0.95 |
| ILCCO (McKay et al. 2017) | 0.44 | 1.69E-05 | 0.95 |
| European meta-analysis | 0.43 | 1.13E-07 | 0.95 |
| European meta-analysis cond. smoking | 0.43 | 0.001 | 0.96 |
| OncoArray European (replication) | 0.42 | 0.051 | 0.97 |
| MVP African | 0.47 | 0.028 | 0.93 |
| OncoArray African (replication) | 0.47 | 0.733 | 0.99 |
| Multi-ancestry meta-analysis | 0.43 | **1.12E-08** | 0.95 |
| OncoArray multi-ancestry (replication) | 0.43 | 0.053 | 0.97 |
| Combined meta-analysis (Multi-ancestry meta-analysis + OncoArray multi-ancestry) | 0.43 | 3.69E-09 | 0.96 |

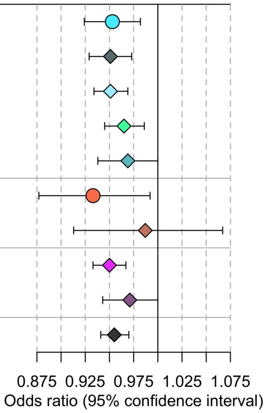


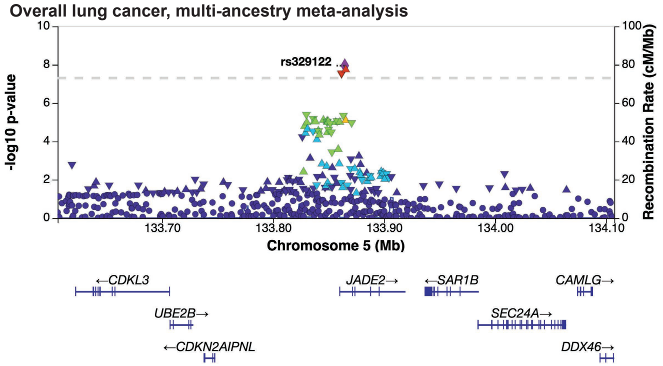


**i**

#### Overall lung cancer, rs7300571 (RPAP3; 12q13.11) : Novel and replicated

| Cohort | EA freq. | P-value | Odds ratio |
|---|---|---|---|
| MVP European | 0.11 | 2.79E-04 | 1.09 |
| ILCCO (McKay et al. 2017) | 0.11 | 8.49E-05 | 1.08 |
| European meta-analysis | 0.11 | 9.74E-08 | 1.08 |
| European meta-analysis cond. smoking | 0.11 | 0.004 | 1.05 |
| OncoArray European (replication) | 0.12 | 0.002 | 1.08 |
| MVP African | 0.02 | 0.074 | 1.23 |
| OncoArray African (replication) | 0.04 | 0.592 | 0.94 |
| Multi-ancestry meta-analysis | 0.11 | **3.47E-08** | 1.08 |
| OncoArray multi-ancestry (replication) | 0.10 | 0.004 | 1.07 |
| Combined meta-analysis (Multi-ancestry meta-analysis + OncoArray multi-ancestry) | 0.11 | 6.48E-10 | 1.08 |

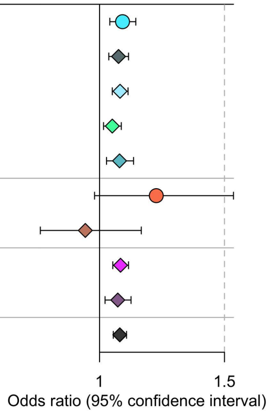


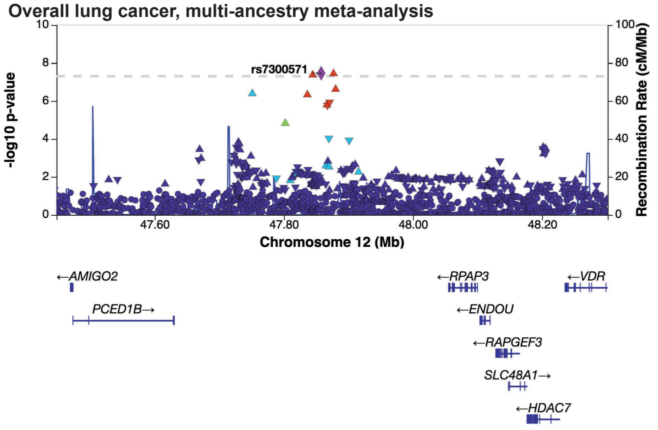

**Supplementary Fig. 4. Genome-wide significant novel lung cancer loci.** Forest plots (left) and regional Manhattan plots (right) for novel loci from European meta-analysis: **a)** *XCL2*, **b)** *LSAMP*, **c)** *NMUR2*, **d)** *TUPL3*, **e)** MYC, **f)** *TLE3*, and **g)** *BLOC1S2*; and from multi-ancestry meta-analysis: **h)** *JADE2*; **i)** *RPAP3*. Manhattan plot SNP data triangles pointing upwards or downwards correspond to positive or negative effect direction, based on the effect allele defined in summary statistics.

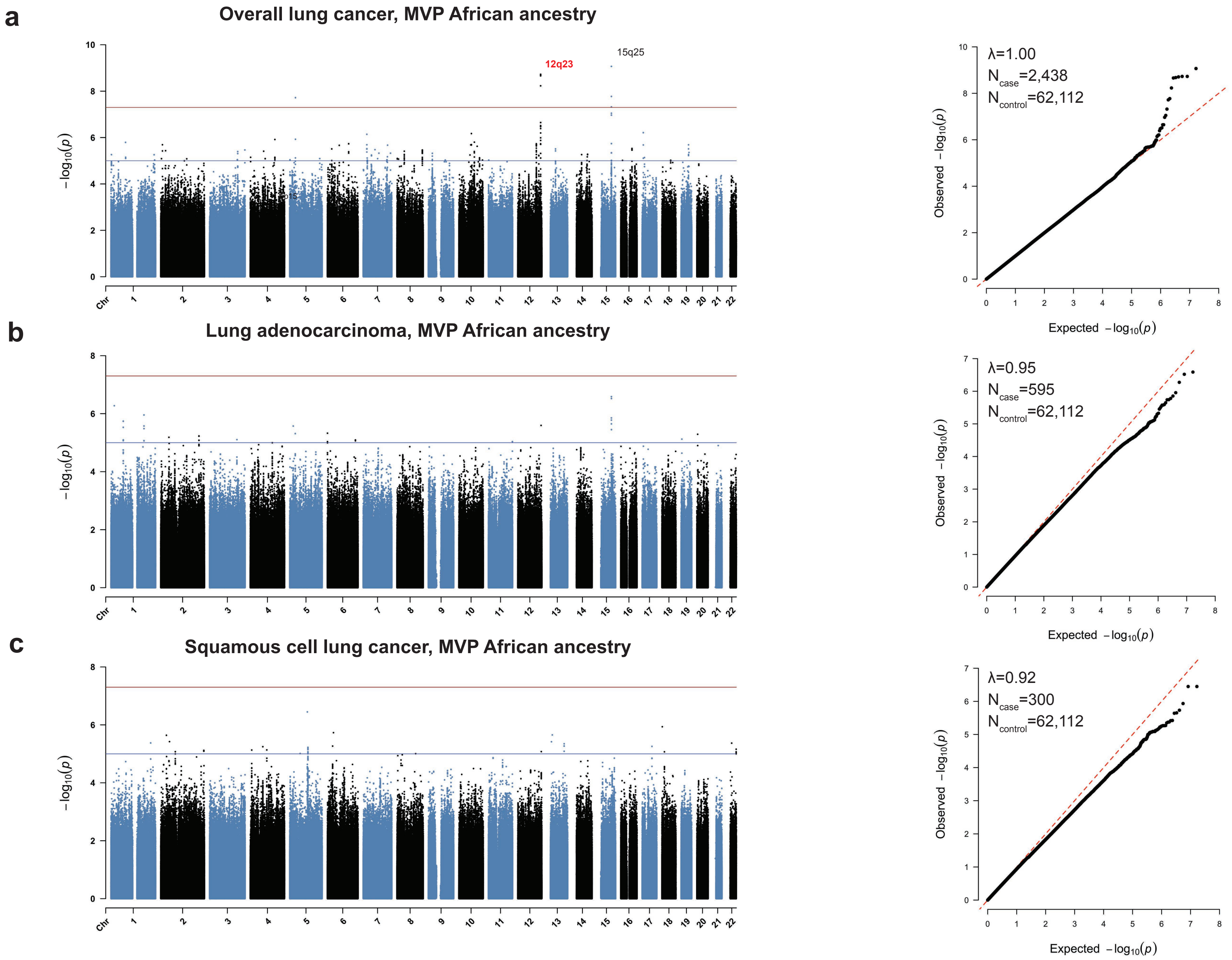


**Supplementary Fig. 5. Manhattan plots and quantile-quantile (QQ) plots for MVP African ancestry.** Manhattan and QQ plots are shown for **a)** African ancestry overall lung cancer; **b)** lung adenocarcinoma (LUAD); and **c)** squamous cell lung carcinoma (LUSC). Cytoband positions for significant loci are noted in each Manhattan plot; putatively novel loci identified in this study are in red. Genomic control (λ) values and sample sizes are inset in QQ plots.

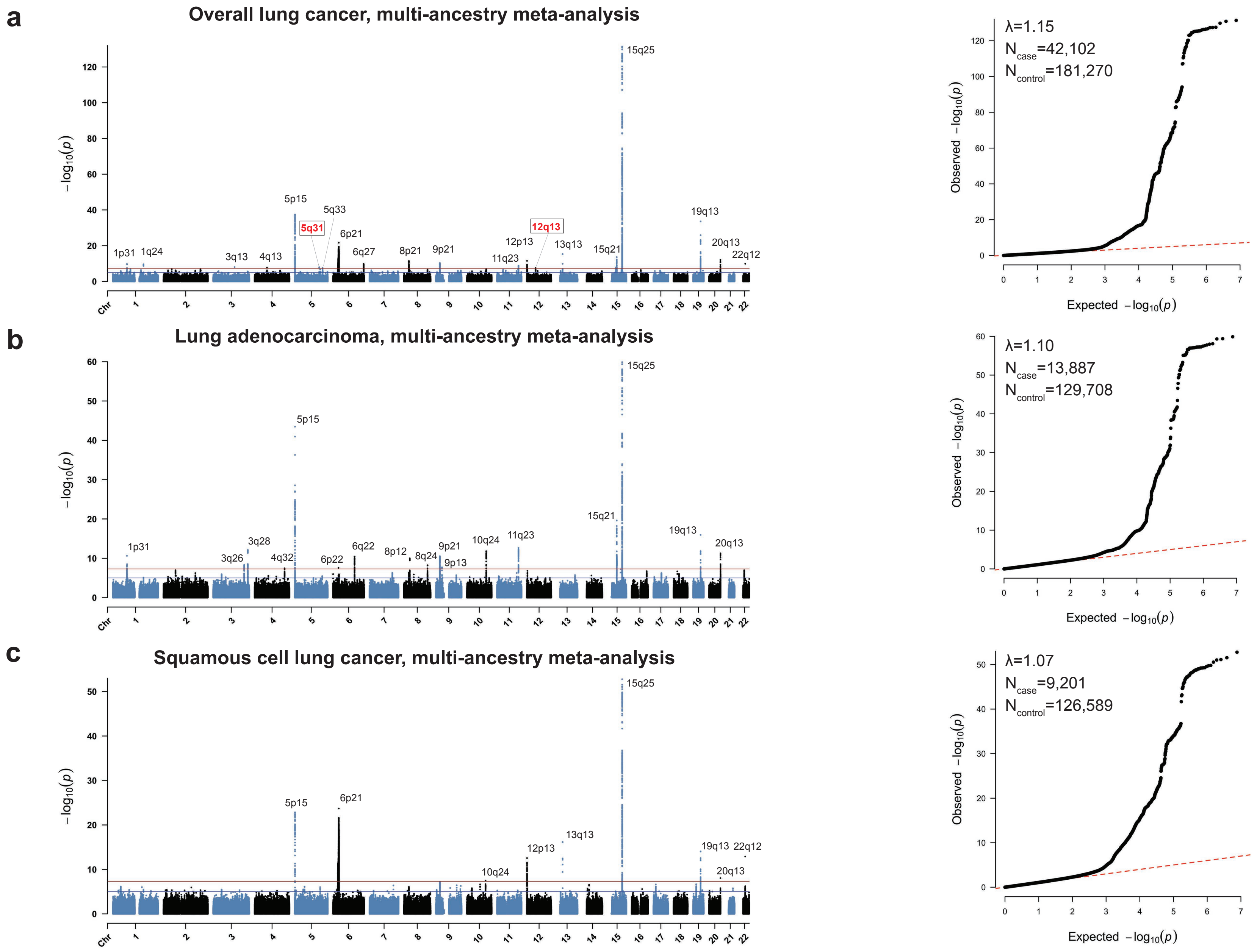


**Supplementary Fig. 6. Manhattan plots and quantile-quantile (QQ) plots for multi-ancestry meta-analyses.** Manhattan and QQ plots are shown for **a)** the multi-ancestry meta-analysis in overall lung cancer; **b)** lung adenocarcinoma (LUAD); and **c)** squamous cell lung carcinoma (LUSC). Cytoband positions for significant loci are noted in each Manhattan plot; novel loci not identified in the European meta-analysis are in red; externally replicated novel loci are indicated by a box. Genomic control (λ) values and sample sizes are inset in QQ plots.

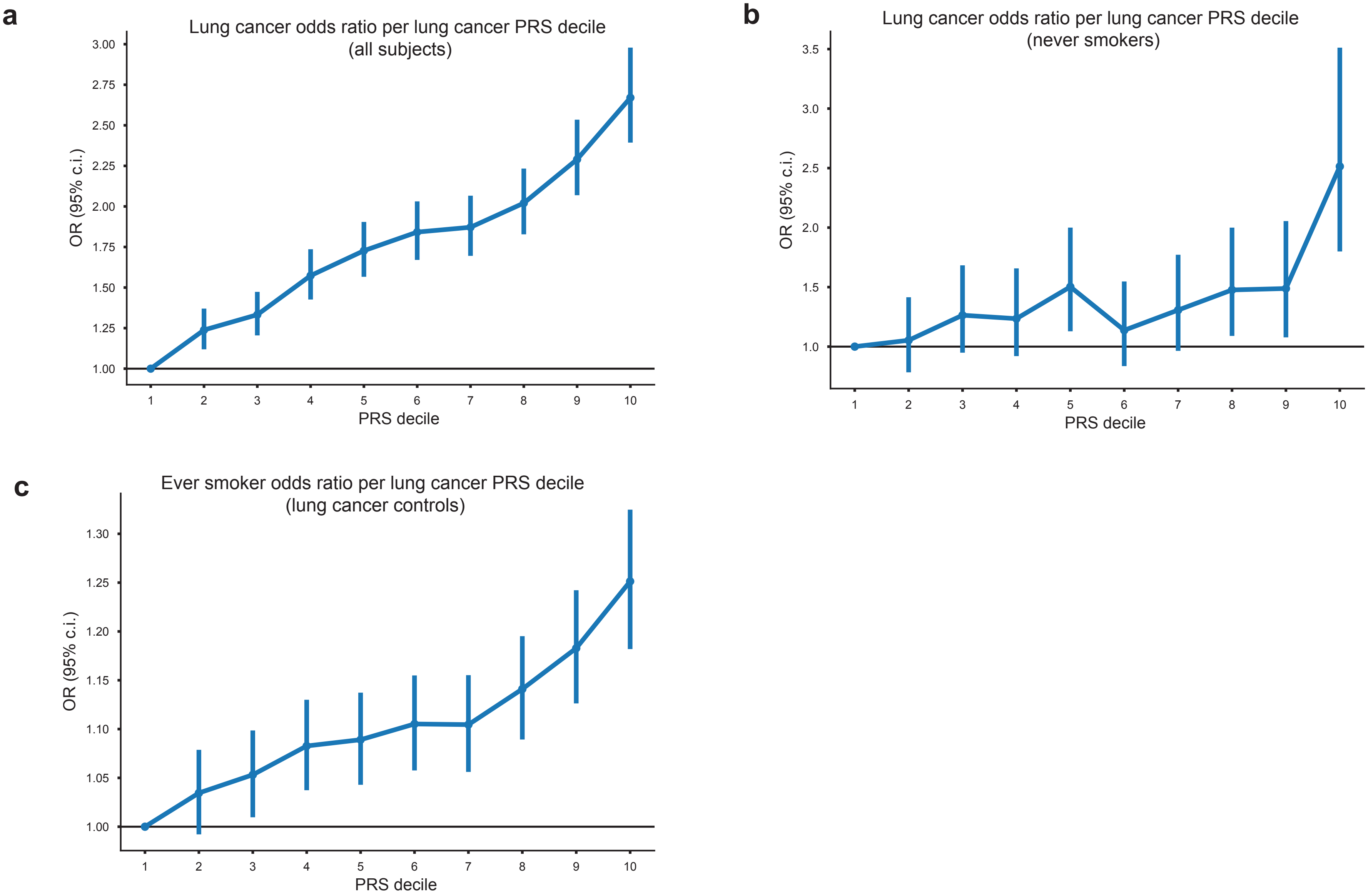


**Supplementary Fig. 7. Association of the lung cancer polygenic risk score (PRS) with lung cancer by smoking status. a)** Association of the lung cancer PRS with overall lung cancer risk. The risk of lung cancer reached an odds ratio (OR) of 2.51 (95% confidence interval: 1.80, 3.51) in the top decile. **b)** Association of the lung cancer PRS with lung cancer risk in never-smokers. Among never-smokers, lung cancer risk reached an OR of 2.67 (2.40, 2.98) in the top decile. **c)** Association of the lung cancer PRS with lung cancer risk in ever-smokers with no history of lung cancer. The top PRS decile was associated with an OR of 1.25 (1.18, 1.32).

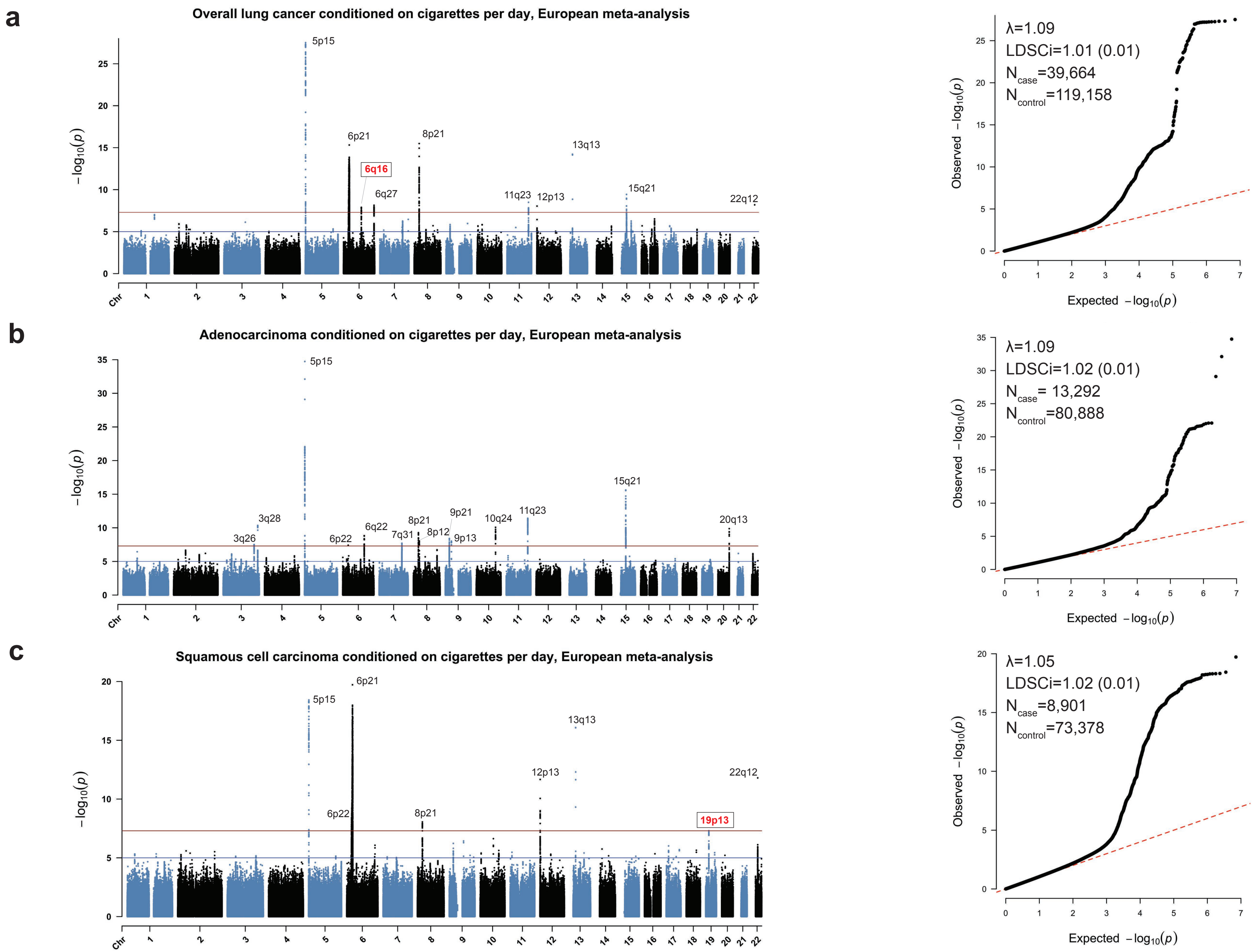


**Supplementary Fig. 8. Manhattan plots and quantile-quantile (QQ) plots for European meta-analyses conditioned on cigarettes per day**
Manhattan and QQ plots for **a)** overall lung cancer conditioned on cigarettes per day; **b)** lung adenocarcinoma (LUAD) conditioned on cigarettes per day; and **c)** squamous cell lung carcinoma (LUSC) conditioned on cigarettes per day. Cytoband positions for significant loci are noted in each Manhattan plot; novel loci not identified in the European meta-analysis are in red; externally replicated novel loci are indicated by a box. Genomic control (λ) values, LDSC intercepts, and sample sizes are inset in QQ plots.

**S9a** Overall lung cancer, rs1124241 (MMS22L; 6q16.1) : Novel and replicated

| Cohort | EA freq. | P-value | Odds ratio |
|---|---|---|---|
| MVP European | 0.21 | 0.061 | 1.03 |
| ILCCO (McKay et al. 2017) | 0.23 | 1.74E-04 | 1.05 |
| European meta-analysis | 0.22 | 3.84E-05 | 1.05 |
| European meta-analysis cond. smoking | 0.22 | **1.26E-08** | 1.08 |
| OncoArray European (replication) | 0.22 | 0.006 | 1.06 |
| MVP African | 0.21 | 0.964 | 1.00 |
| Multi-ancestry meta-analysis | 0.22 | 7.48E-05 | 1.04 |
| Combined meta-analysis (European meta-analysis cond. smoking + OncoArray European) | 0.22 | 3.39E-10 | 1.07 |

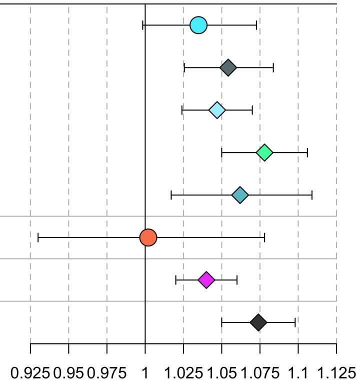


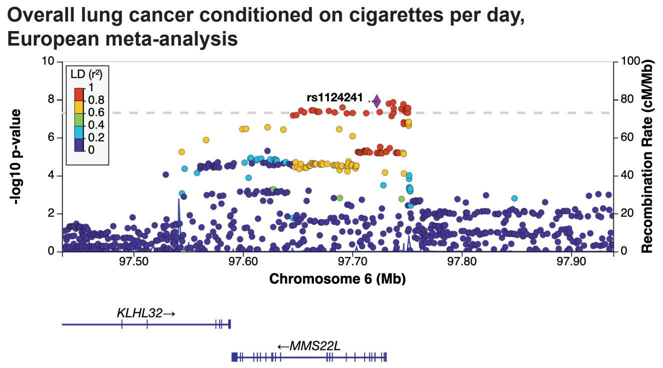


**b** Squamous cell lung carcinoma, rs61494113 (ABHD8; 19p13.11) : Novel and replicated

| Cohort | EA freq. | P-value | Odds ratio |
|---|---|---|---|
| MVP European | 0.29 | 0.027 | 1.10 |
| ILCCO (McKay et al. 2017) | 0.29 | 2.05E-05 | 1.09 |
| European meta-analysis | 0.29 | 1.62E-06 | 1.09 |
| European meta-analysis cond. smoking | 0.29 | **4.90E-08** | 1.12 |
| OncoArray European (replication) | 0.29 | 0.003 | 1.10 |
| MVP African | 0.40 | 0.019 | 1.23 |
| Multi-ancestry meta-analysis | 0.30 | 2.18E-07 | 1.10 |
| Combined meta-analysis (European meta-analysis cond. smoking + OncoArray European) | 0.29 | 6.39E-10 | 1.11 |

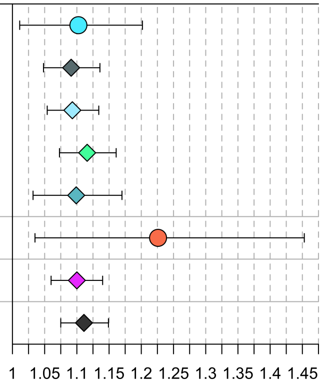


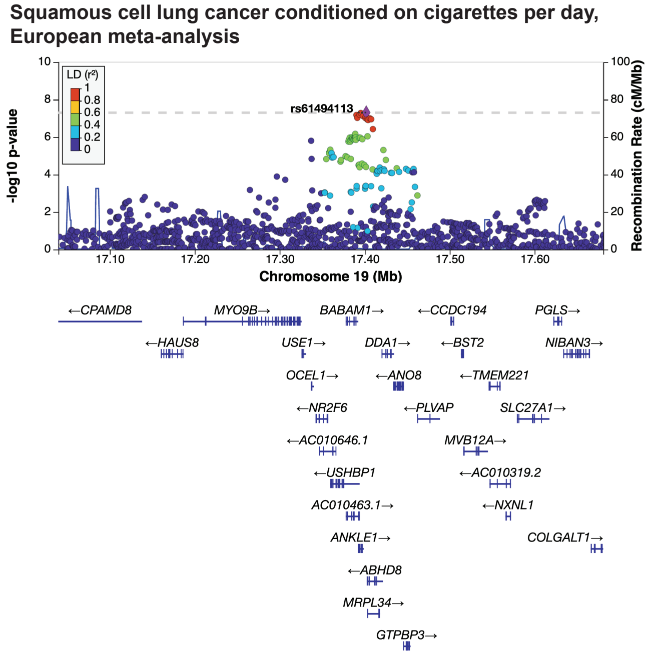


**Supplementary Fig. 9. Novel loci for overall lung cancer and squamous cell carcinoma conditioned on smoking.** Locus zoom plots for the novel loci in the European meta-analysis conditioned on cigarettes per day: **a)** 6q16 in overall lung cancer and **b)** 19p13 in squamous cell lung cancer.
